# Resource requirements for reintroducing elective surgery in England during the COVID-19 pandemic: a modelling study

**DOI:** 10.1101/2020.06.10.20127266

**Authors:** A. J. Fowler, T. D. Dobbs, Y. I. Wan, R. Laloo, S. Hui, D. Nepogodiev, A. Bhangu, I. S. Whitaker, R. M. Pearse, T. E. F. Abbott

## Abstract

**Background:** The COVID-19 response required the cancellation of all but the most urgent surgical procedures. The number of cancelled surgical procedures in the National Health Service (NHS) England due to COVID-19, and the reintroduction of surgical activity, was modelled.

**Methods:** Modelling study using Hospital Episode Statistics data (2014-2019). Using NHS England definitions, surgical procedures were grouped into four urgency classes. Expected numbers of surgical procedures performed between 1^st^ March 2020 and 28^th^ February 2021 were modelled. Procedure deficit was estimated using conservative assumptions and the gradual reintroduction of elective surgery from the 1^st^ June 2020. Costs were calculated using NHS reference costs and are reported as millions(M) or billions(B) of Euros(€). Estimates are reported with 95% confidence intervals.

**Results:** 4 547 534 (3 318 195 – 6 250 771) patients with pooled mean age of 53.5 years were expected to undergo surgery between 1^st^ March 2020 and 28^th^ February 2021. By 31^st^ May 2020, 749 247 (513 564 – 1 077 448) surgical procedures were cancelled. Assuming elective surgery is gradually reintroduced, 2 328 193 (1 483 834 – 3 450 043) patients will be awaiting surgery by 28th February 2021. The cost of delayed procedures is €5.3B (€3.1B - €8.0B). Safe delivery of surgery during the pandemic will require substantial extra resources costing €526.8M (€449.6M - €633.9M).

**Conclusion:** Reintroduction of elective surgery in NHS England will be associated with substantial treatment delays, and large cost increases. The challenges and costs of reintroducing surgical care in other healthcare settings may differ and further research to monitor the recovery of surgical care is urgently required.

## Introduction

The outbreak of novel coronavirus SARS-CoV-2 and the disease COVID-19, were first reported in December 2019 in Wuhan, China^1^ and the World Health Organisation declared a global pandemic in March 2020. The healthcare response to the COVID-19 pandemic has required rapid changes to the provision of secondary healthcare services, including the creation of new hospitals.^2, 3^ Critical care capacity has increased substantially in high income countries, requiring re-deployment of staff and equipment from other departments.^4^ Surgical services represent a large portion of healthcare activity, accounting for 5 million hospital admissions to the United Kingdom (UK) National Health Service (NHS) every year.^5, 6^ In March, National Health Service (NHS) England issued guidance to postpone non-urgent surgery from 15^th^ April.^7^ Only very urgent and emergency surgical treatments continued, with the cancellation of almost all planned cancer and non-cancer surgery.^8, 9^ However, the number of surgical procedures cancelled during the pandemic response, and the excess of untreated surgical disease, remain unknown.^2, 8^

As the NHS response to the pandemic develops, there is a planned reintroduction of elective surgery with a particular focus on cancer surgery.^10^ However, the risk of SARS-CoV-2 transmission within hospitals is a concern and further complicated by uncertain rates of asymptomatic infection in the general population (17% to 56%).^11, 12^ Asymptomatic infection rates may be as high as 24% among healthcare staff.^13, 14^ New procedures introduced to protect patients and staff from virus transmission have reduced capacity for surgery, as has a lack of appropriately trained staff requiring redeployment to other clinical areas.^2, 4^ NHS plans to prevent in-hospital infection include preoperative patient screening, establishing ‘clean’ hospitals, routine staff testing and/or universal personal protective equipment (PPE).^8, 15^ However, the optimum strategy, resource requirements and time-scale for reintroducing elective surgery remain uncertain.^15^ Critical care utilisation remained high for patients with COVID-19 in some areas after the peak passed.^16^ Capacity for surgical care remains limited in operating rooms, post-anaesthetic recovery units and critical care units whilst the incidence of serious postoperative complications amongst patients with COVID-19 is very high.^4, 17^ A clear understanding of the number and type of overdue surgical procedures is urgently needed to inform national healthcare policies for reintroducing surgical services.

Hospital Episode Statistics (HES) data over five years from 2014 to 2019 were used to estimate the number of surgical procedures postponed in the NHS in England due to COVID-19. Multiple scenarios for the reintroduction of routine surgical activity over nine months, starting in June 2020, were modelled. Estimates of the cumulative national deficit of surgical procedures and the resource implications for ‘catching up’ with postponed procedures, in terms of hospital beds, critical care beds, PPE for staff, preoperative screening of patients for COVID-19, and the associated financial cost are presented.

## Methods

An unabridged description of the methods is provided in the appendix.

### Data source

Aggregated HES data for Admitted Patient Care (APC) from between 1^st^ April 2014 and 31^st^ March 2019 (https://digital.nhs.uk/data-and-information) were used. HES provide detailed data describing hospital care in England, including the type of surgical procedure categorised by an Office for Population Censuses Surveys Classification of Interventions and Procedures (OPCS) codes. All data used are freely available anonymised data and research ethics and information governance approvals were therefore not required.

### Outcomes

The primary outcome measure was the deficit of surgical activity, defined as the number of cancelled surgical procedures, attributable to the COVID-19 pandemic response. The secondary outcomes were the associated resource requirements, defined as: hospital bed days, number of critical care admissions, number of investigations for preoperative COVID-19 screening, the amount of PPE required for operating theatre staff and the additional financial cost.

### Modelling analysis

A statistical analysis plan was developed and published before data analysis took place.^18^ R version 3.6.1 (R Core Team, Vienna, Austria) was used for data analysis. Surgical procedures were stratified according to a classification published by NHS England on 17^th^ March 2020, which described operations that should be stopped or continued during the initial pandemic response.^7^ Surgical procedures were divided into four classes of decreasing urgency: Class 1 – Emergency operations needed within 72 hours; Class 2 – Urgent surgery that can be deferred for up to 4 weeks; Class 3 – Semi-urgent surgery that can be delayed for up to 3 months; and Class 4 – Elective surgery that can be delayed for more than 3 months. A pre-specified, data-driven approach was used to identify emergency, urgent and elective surgical procedures based on waiting times and expert review of procedure coding (supplementary tables 1 and 2). The monthly volume of surgical activity that would have been expected had there not been a pandemic until 28^th^ February 2021 was estimated, by calculating the annual change in activity during the five-year period from 1^st^ April 2014 to 31^st^ March 2019, and extrapolated using a linear growth assumption. Estimations of monthly volume were calculated using NHS England Monthly Activity Returns, and the average age for each class of surgery was estimated.

### Surgical procedure volume

The number of postponed or cancelled surgical procedures from the 1^st^ March 2020 up to 1^st^ June 2020 were estimated according to several assumptions. First, that class 1 (emergency) surgery would continue at the pre-pandemic rate. Second, that class 2 surgery would continue at a reduced rate. Four scenarios were calculated, where 20%, 40%, 60% and 80% of class 2 surgical procedures were assumed to have continued. Third, that 50% of class 3 and 4 procedures continued in March and then stopped completely in April and May, reflecting between-hospital heterogeneity in the timing of stoppage of surgery. The results were presented as the deficit of surgical procedures between 1^st^ March and 31^st^ May 2020 with a 95% confidence interval. Fourth, that widespread reintroduction of surgical activity would start from 1^st^ June 2020 and continue to increase to pre-pandemic levels. For each of the four scenarios of class 2 procedures, a linear increase in activity over the three months from 1^st^ June to 31^st^ August 2020 was assumed, with class 3 and 4 surgical procedures remaining cancelled until 31^st^ August. In the final model it was assumed that 80% of class 2 procedures continued. This analysis was repeated in an iterative fashion, by adding class 3 procedures on 1^st^ September and class 4 procedures on 1^st^ December. Fifth, that pre-pandemic levels of surgical activity would be reached by 28^th^ February 2021 (supplementary figure 1). The estimated number of surgical procedures carried out each month between 1^st^ March 2020 and 28^th^ February 2021 and a rolling deficit of surgical activity compared to the expected volume of surgery according to the previous five-year average are presented. Assumptions about a second peak of COVID-19 were not included, and neither were assumptions regarding the impact of reduced operating theatre utilisation due to enhanced infection control procedures. A post-hoc sensitivity analysis was performed assuming that all classes of surgery restarted one the 1^st^ June 2020 and increased linearly over a 6-month period.

### Hospital admissions

The total number of bed-days, weighted by procedure frequency, were calculated by multiplying the median length of stay by the number of inpatient admissions. The proportion of patients that would require postoperative critical care were estimated using a conservative assumption of 1% of patients undergoing inpatient surgery, and 4% of those undergoing emergency surgery.^19-21^

### Preoperative screening tests

Preoperative COVID-19 screening requirements were modelled according to three scenarios. First, all patients would have two outpatient preoperative COVID-19 Polymerase Chain Reaction (PCR) tests. Second, all patients would have one outpatient and one inpatient preoperative SARS-CoV-2 PCR tests, requiring an additional day in hospital for isolation. Third, all patients would have one outpatient and one inpatient preoperative SARS-CoV-2 PCR tests, and patients undergoing class 1 (emergency) thoracic, cardiac or abdominal procedures would also have a computed tomography scan of the chest, in line with guidance from the Royal College of Radiologists.^22, 23^

### Personal protective equipment

The amount of PPE required in operating theatres was estimated according to two scenarios.^24^ First, that eight staff members would be present for every procedure (two surgeons, two anaesthetists, one operating department practitioner and three scrub staff). Second, that four staff members would be present for every procedure (one surgeon, one anaesthetist, one operating department practitioner and one scrub staff), each requiring an FFP3 mask, a fluid repellent gown, two pairs of gloves and eye protection.

### Estimated financial cost

The estimated financial cost of reintroducing surgical activity was divided into three areas: the cost of the surgical procedure; the cost of preoperative COVID-19 screening arrangements; and, the cost of PPE required. The total cost of reintroducing surgical activity from 1^st^ June 2020 and the total deficit of surgical procedures on 28^th^ February 2021 was estimated by combining these. Costs were calculated in Great British Pounds (£) and are presented in Euros (€) based on the average exchange rate reported by OANDA on the 1^st^ July 2020 (£1 = €1.09766). Costs of millions (M) and billions (B) are reported as whole amounts, rounded to one decimal place. The total costs of surgical procedures was calculated by matching the OPCS v4.7 code with the Health Resource Group coding and summing the associated procedure cost according to the national schedule of NHS costs in 2015.^5, 25^ The costs of screening tests were calculated according to £19 (€20.86) per SARS-CoV-2 PCR and £69 (€75.74) per CT scan and a range of between £222 (€243.68) and £346 (€379.79) per additional bed day.^26-28^ The costs of PPE was calculated as £2.90 (€3.18) per FFP3 mask, £14.90 (€16.36) per 100 gloves, £3 (€3.29) per fluid resistant gown and £2.90 (€3.18) per piece of eye protection.^28, 29^

## Results

Some 1073 OPCS codes for Class 1 – 4 surgical procedures, representing a total of 22 513 872 surgical admissions between 1^st^ April 2014 and 31^st^ March 2019 (Figure 1). The monthly median number of procedures was 382 768 (Interquartile range [IQR]: 22 890) (supplementary table 3). If growth in the number of surgical procedures had continued according to a pre-pandemic trajectory, 4 547 534 (95%CI: 3 318 195 to 6 250 771) would have been performed between 1^st^ March 2020 and 28^th^ February 2021 (supplementary table 4). Patients aged over 60 years accounted for 32.7% of class 1, 43.9% of class 2, 53.7% of class 3 and 45.6% of class 4 surgical activity (supplementary table 5).

### Surgical procedure volume

Between 1^st^ March and 31^st^ May 2020, 749 247 (95%CI: 513 564 to 1 077 448) surgical procedures will have been cancelled in comparison with observed pre-pandemic levels of activity (supplementary table 6). The widespread reintroduction of class 2 surgical activity from 31^st^ May 2020, was modelled for four scenarios of incrementally increasing activity (20% - 80%) that would reach pre-pandemic levels by 31^st^ August 2020 (supplementary table 7). Even if class 2 - 4 surgical procedures are reintroduced in a stepwise fashion between 1^st^ June 2020 and 28^th^ February 2021 to reach the predicted pre-pandemic level of activity, the number of cancelled surgical procedures would be 2 328 193 (95% CI: 1 483 834 to 3 450 043) procedures (table 1, figures 2-3). In a post-hoc sensitivity analysis assuming all classes of surgery restarted on 1^st^ June and took six months to get to normal capacity, the number of cancelled surgical procedures was estimated to be 1 551 560 (95%CI: 1 041 537 to 2 269 486) (supplementary table 11).

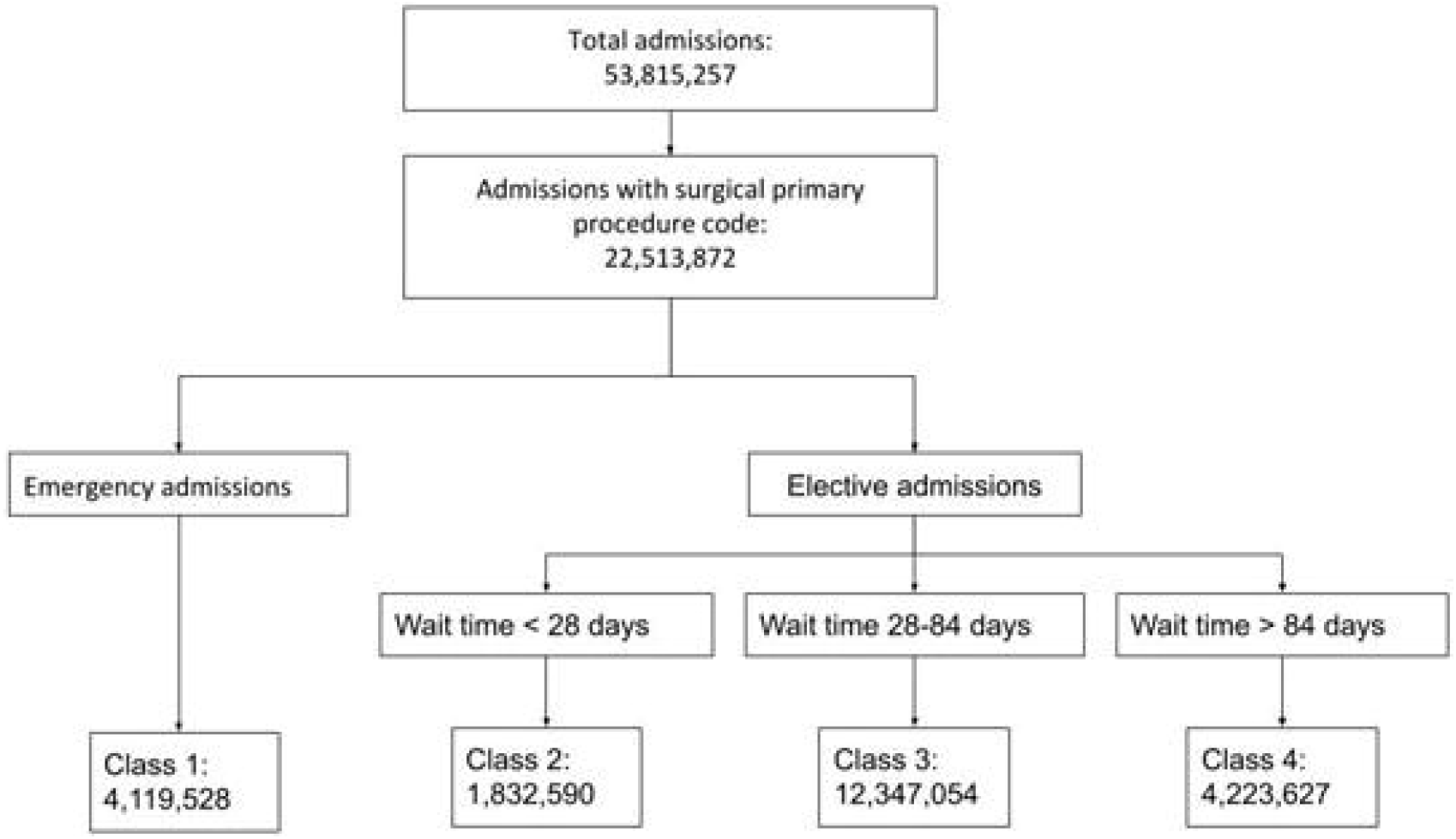

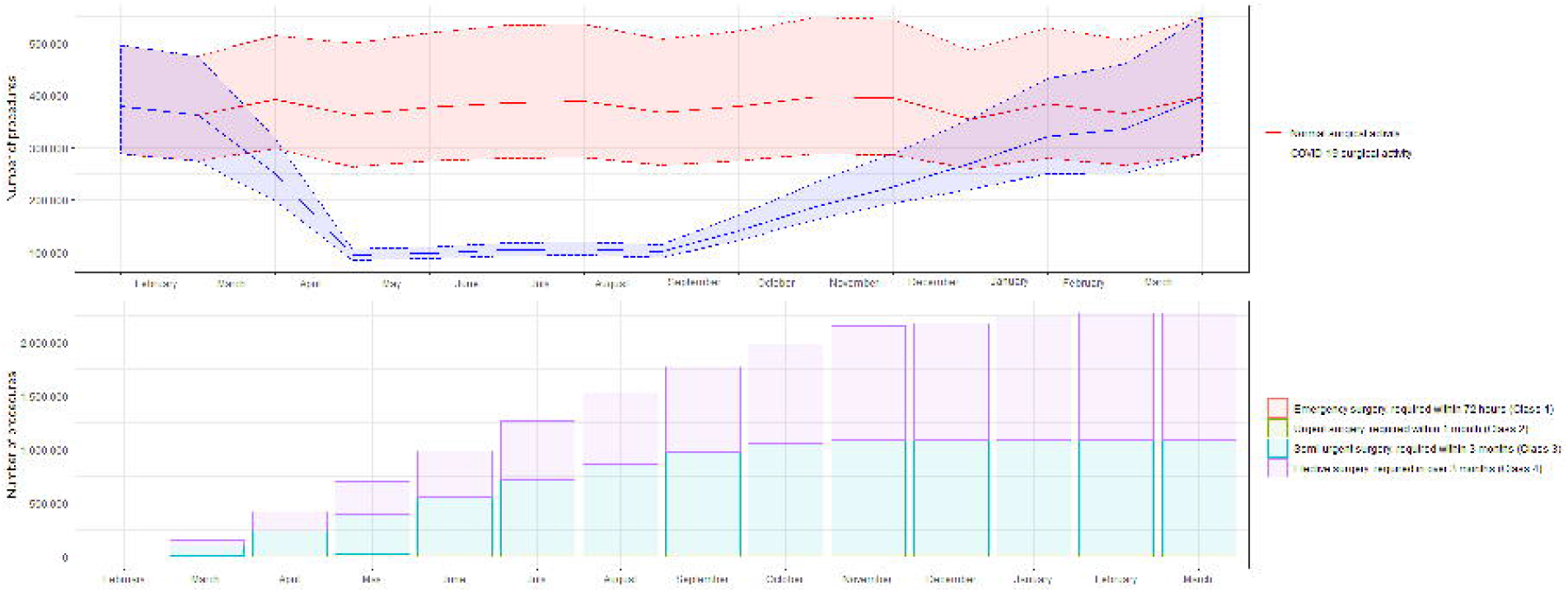

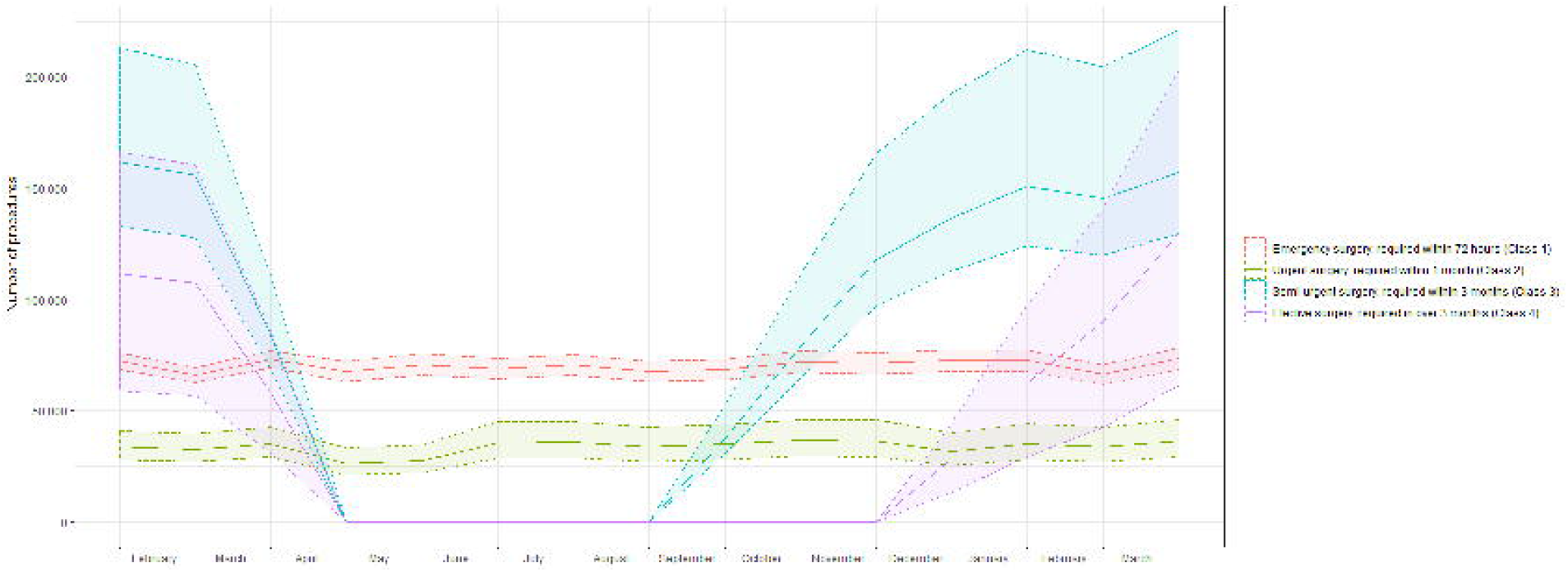

### Hospital admissions

The total bed days associated with the deficit of surgical activity on 31^st^ May 2020 is 973 006 (95%CI: 623 700 to 1 423 014) days (supplementary table 8). If widespread reintroduction of surgical activity occurs from 1^st^ June 2020, the total number of bed days associated with the cumulative deficit of surgical activity up to 28^th^ February 2021 is 3 337 706 (95%CI: 1 997 510 to 4 995 117) days (supplementary table 8). The total number of critical care admissions associated with the deficit of surgical activity on 31^st^ May 2020 is 2 474 (95%CI: 1 536 to 3 650) (supplementary table 8). If widespread reintroduction of surgical activity occurs from 1^st^ June 2020, the estimated total number of critical care admissions associated with the cumulative deficit of surgery up to 28th February 2021 will be 8 769 admissions (95%CI: 5 103 to 13 207) (supplementary table 8).

### Preoperative screening tests

The estimated resource requirement for preoperative COVID-19 screening associated with the deficit of surgical activity on 31^st^ May 2020 include: 1 390 104 (95%CI: 939 958 to 2 018 502) SARS-CoV-2 PCR tests and 247 321 (95%CI: 153 633 to 365 080) bed days for preoperative isolation. The screening resource requirements associated with the reintroduction of surgical activity from 1^st^ June 2020 is provided in table 2.

### Personal protective equipment

The estimated amount of PPE associated with the deficit of surgical activity on 31^st^ May 2020 is 11 120 832 (95% CI: 7 519 664 to 16 148 016) items, assuming four persons per theatre, and 22 241 664 (95% CI: 15 039 328 to 32 296 032) items assuming eight persons pre theatre. The estimated total amount of PPE associated with the cumulative deficit of surgical activity by 28th February 2021 is between 37 251 088 (95%CI: 23 741 344 to 55 200 672) items, assuming four persons per theatre and 74 502 176 items (95%CI: 47 482 688 to 110 401 344) assuming eight persons per theatre (supplementary table 8).

### Financial cost

The procedure cost for the deficit of surgical activity on 31^st^ May 2020 is €1.5B (95%CI: €923.0M to €2.2B) (table 2 and supplementary table 10). If elective surgery recommences between 1^st^ June 2020 and 28^th^ February, the total costs associated with performing reintroduced surgery, including the costs of the procedure, PPE and screening, are between €5.4B (95%CI: €4.4B to €6.7B) and €5.7B (95%CI: €4.7B to €7.0B) (supplementary table 9). The total cost of cancelled operations by 28^th^ February 2021, including the costs of the procedure, PPE and screening tests, are between €5.7B (95%CI: €3.3B to €8.6B) and €5.9B (95%CI: €3.5B to €8.9B) (supplementary table 10).

An unabridged description of the results is provided in the appendix.

## Discussion

The principal finding of this study is that, without a substantial increase in capacity, accumulated delays in surgical care will lead to a backlog of more than two million overdue or cancelled surgical procedures in the NHS in England by 28^th^ February 2021. This is equivalent to 45% of the total number of surgical procedures performed in England each year before the COVID-19 pandemic, and is larger than the existing waiting list to start surgical treatment in March 2020, which already stood at over 1.8 million patients.^30^ The cost of clearing this post-pandemic waiting list is in excess of €5 billion, or 4.2% of the total NHS England budget.^31^ Additional costs of delivering surgical services under strict infection control procedures exceed €500 million, and include personal protective equipment, preoperative screening and extra bed-days in hospital. No attempt was made to model the reduced operating theatre utilisation efficiency, suggesting the true excess cost may be far greater. Nor does this estimate account for additional costs of contracts with non-NHS providers, or the costs of postoperative complications among surgical patients with COVID-19.^4, 17^ In order to reduce the size of any future backlog of overdue surgery, elective surgical services would need to be reintroduced at the earliest opportunity and at the maximum capacity possible. Whether this can be achieved safely remains unclear.^15^

Postoperative mortality is much higher in patients with COVID-19 infection, compared to the expected pre-pandemic rate.^17^ An international cohort study of more than 1000 patients with COVID-19 found the incidence of postoperative mortality was more than one in four patients, compared to an expected mortality rate of 1 in 30 patients undergoing emergency surgery.^17^ This will inevitably lead to higher resource use, longer in-patient lengths of stay and greater financial costs. Furthermore, emerging data from the UK suggests that as many as one in twelve healthcare workers had asymptomatic infection during the first peak of the pandemic, and were at risk of transmitting the virus to other staff and patients.^14^ Consequently, many surgeons may choose to avoid surgery in patients with COVID-19 infection, to reduce the active reservoir inside the hospital and to mitigate the higher-risk of postoperative mortality. When surgery does go ahead, the benefits must be weighed against the potential harm of bringing vulnerable patients into contact with SARS-CoV-2 and the potential risks to staff members. It is likely that preoperative screening for the virus and PPE for healthcare workers will become universal. It is estimated that by February 2021, over 160 000 SARS-CoV-2 PCR tests will be required every week for preoperative screening alone. Permanent infrastructure for testing will need to be included in strategic plans for pathology services to support elective surgery. There are no national data regarding the use of protective equipment at individual institutions. However, many societies have exercised caution and have advocated for the deployment of full PPE for all high risk procedures and examinations, including all aerosol generating procedures.^24^

There are no published data on the rate of cancellation of surgery within the NHS in England during the COVID-19 pandemic response, of which we are aware. However, the results of this analysis are consistent with data from an international survey, which reported that the majority of both urgent cancer and non-cancer surgery were postponed during the peak of the pandemic.^9^ This analysis provides early evidence that disruption to surgical care has been widespread and reintroduction of this care will have substantial resource requirements that will differ between healthcare settings. Delays to the re-introduction of surgical procedures may result in increased secondary morbidity and mortality, not directly attributable to COVID-19 infection, but due to increased volume of undiagnosed surgical disease and delays to curative therapy.^2^ If delays in surgical care result in higher incidence of more advanced disease, it is likely that postoperative morbidity and mortality will also increase, as well as critical care bed utilisation and hospital length of stay. Strategies for undertaking safe surgery after the pandemic, during a prolonged pandemic response, or if COVID-19 becomes endemic, include reorganisation of referral pathways, restructuring of the surgical workforce, dedicated surgical critical care resources, isolated ‘cold’ hospitals and prioritisation of certain patient groups.^2^ If second or subsequent peaks of COVID-19 occur, it is likely surgical services will again be curtailed, with further increases in the number of cancelled procedures. NHS leaders should make plans for the future continuity of surgical care.

This analysis has several strengths. Robust national data were analysed that describe the whole healthcare setting of a single country according to a prospectively published statistical analysis plan. Conservative estimates are purposely provided of estimates of procedure volume and associated resource requirements. Procedures are reported at the admission level, rather than the episode level, which will likely underestimate the volume of surgery. Assignment to urgency class based on mean wait time may lead to some inappropriate classification of individual procedures but was the only feasible option. Multiple sources were used to calculate postoperative critical care admission, which did not include preoperative admission, nor increased critical care utilisation for patients with COVID-19. Estimates of PPE only include usage in operating theatre, but not on postoperative wards or in clinics. Cost estimates do not take account of potential future reorganisation of care pathways, the use of ‘hot’ and ‘cold’ sites or use of private sector hospitals. This analysis also has limitations. A series of assumptions were made based on NHS England guidance, which were applied to a series of plausible scenarios regarding the continuation of surgical activity during the pandemic.^7, 10^ However, the reality of the volume, type and timing of continuation and reintroduction of surgery may differ from these models. The models implemented in this analysis are highly sensitive to the rate and timing of the resumption of surgical care, as evidenced by the much lower cumulative deficit in the sensitivity analysis. It will only be possible to test this in retrospect once actual numbers of procedures carried out are published. Our analysis does not account for lower throughput of surgery due to stricter infection control procedures or a potential second peak of COVID-19.^32^ Further research is urgently needed to address these issues. It is likely that the volume of emergency surgery that has continued during the pandemic is lower than pre-pandemic levels, perhaps due to patients avoiding hospital or clinicians using alternative management strategies.^33^ The age distribution of the surgical population is skewed.^6^ However, due to limitations of the data source, age adjustment in the analysis was not possible. It is likely that, unfortunately, some patients waiting for surgery will have died while waiting to have their surgery. The numbers of these cases are unknown, and it was not possible to account for these in the analysis. It is likely that patients who have delayed care will have higher care needs and associated financial costs. There will also be higher human costs in terms of chronic symptoms and disability that it was not possible to account for in this analysis.^2^ This analysis will be updated as new data become available.

In conclusion, accumulated delays in surgical care due to COVID-19 will lead to a backlog of more than two million overdue or cancelled surgical procedures in the NHS in England by 28^th^ February 2021. These delays in care are likely to be associated with increased mortality. The procedure cost of clearing this waiting list is more than €5.3 billion, with additional costs of at least €400 million for personal protective equipment and preoperative screening. Further research is needed to provide regular reports of NHS surgical activity across the United Kingdom, to support strategic planning of the re-introduction of elective surgery nationwide.

## Supporting information

main tables for manuscript

## Data Availability

All data is freely available on the NHS Digital Website and the NHS England Website. The definition of surgery is available in the supplement of our publication.

https://digital.nhs.uk/data-and-information/publications/statistical/hospital-admitted-patient-care-activity

https://www.england.nhs.uk/statistics/statistical-work-areas/hospital-activity/monthly-hospital-activity/mar-data/

https://academic.oup.com/bja/article/119/2/249/4049141

## Contributions

AF, TD and TA were responsible for study design. RL, SH, TD, YW and AF were responsible for data collection. AF and TA were responsible for data analysis. AF, TD, RP and TA were responsible for data interpretation. TA wrote the first draft of the manuscript. All authors revised the manuscript for important intellectual content and approved the final version.

## Data sharing

The data source is freely available online, the analysis code is freely available online at (https://github.com/AJFOWLER/covid_surgical_modelling).

## Preregistration

A statistical analysis plan was published on our website prior to starting the analysis (doi: 10.17636/64678).

## Acknowledgements

AF holds a National Institute for Health Research Doctoral Research fellowship.

## References

1. Chen N, Zhou M, Dong X, et al. Epidemiological and clinical characteristics of 99 cases of 2019 novel coronavirus pneumonia in Wuhan, China: a descriptive study. Lancet 2020; 395(10223): 507–13.

2. Soreide K, Hallet J, Matthews JB, et al. Immediate and long-term impact of the COVID-19 pandemic on delivery of surgical services. The British journal of surgery 2020.

3. Abu Hilal M, Besselink MG, Lemmers DHL, Taylor MA, Triboldi A. Early look at the future of healthcare during the COVID-19 pandemic. The British journal of surgery 2020.

4. Collaborative CO. Global guidance for surgical care during the COVID-19 pandemic. The British journal of surgery 2020.

5. Abbott TEF, Fowler AJ, Dobbs TD, Harrison EM, Gillies MA, Pearse RM. Frequency of surgical treatment and related hospital procedures in the UK: a national ecological study using hospital episode statistics. British journal of anaesthesia 2017; 119(2): 249–57.

6. Fowler AJ, Abbott TEF, Prowle J, Pearse RM. Age of patients undergoing surgery. The British journal of surgery 2019; 106(8): 1012–8.

7. England N. Clinical guide to surgical prioritsation during the coronavirus pandemic, 2020.

8. Spinelli A, Pellino G. COVID-19 pandemic: perspectives on an unfolding crisis. The British journal of surgery 2020.

9. CovidSurg C, Nepogodiev D, Bhangu A. Elective surgery cancellations due to the COVID-19 pandemic: global predictive modelling to inform surgical recovery plans. The British journal of surgery 2020.

10. England N. Important - for action - second phase of NHS response to COVID19. In: trusts CeoaNTaf, editor. NHS England website: NHS England; 2020.

11. Arons MM, Hatfield KM, Reddy SC, et al. Presymptomatic SARS-CoV-2 Infections and Transmission in a Skilled Nursing Facility. The New England journal of medicine 2020.

12. Mizumoto K, Kagaya K, Zarebski A, Chowell G. Estimating the asymptomatic proportion of coronavirus disease 2019 (COVID-19) cases on board the Diamond Princess cruise ship, Yokohama, Japan, 2020. Euro surveillance : bulletin Europeen sur les maladies transmissibles = European communicable disease bulletin 2020; 25(10).

13. Shields A. SARS-CoV-2 seroconversion in health care workers. MedRxiv 2020.

14. Treibel T. COVID-19: PCR screening of asymptomatic health-care workers at London hospital. Lancet 2020.

15. Mayol J, Fernandez Perez C. Elective surgery after the pandemic: waves beyond the horizon. The British journal of surgery 2020.

16. centre ICnaar. ICNARC report on COVID-19 in critical care 08 May 2020. 2020.

17. Collaborative C. Mortality and pulmonary complications in patients undergoing surgery with perioperative SARS-CoV-2 infection: an international cohort study. Lancet 2020.

18. Fowler AJ, Dobbs TD, Wan YI, et al. Estimated surgical requirements in England after COVID-19: a modelling study using hospital episode statistics. MedRxiv 2020; 09/05/2020.

19. International Surgical Outcomes Study g. Global patient outcomes after elective surgery: prospective cohort study in 27 low-, middle- and high-income countries. British journal of anaesthesia 2016; 117(5): 601–9.

20. Pearse RM, Moreno RP, Bauer P, et al. Mortality after surgery in Europe: a 7 day cohort study. Lancet 2012; 380(9847): 1059–65.

21. Centre ICNAaR. Summary Statistics. 2019. https://http://www.icnarc.org/Our-Audit/Audits/Cmp/Reports/Summary-Statistics (accessed 20/05/2020 2020).

22. Lima DS, Ribeiro MAF, Jr., Gallo G, Di Saverio S. Role of chest CT in patients with acute abdomen during the COVID-19 era. The British journal of surgery 2020.

23. Radiologists TRCo. Statement on use of CT chest to screen for COVID-19 in pre-operative patients. 14/05/2020 2020. https://http://www.rcr.ac.uk/college/coronavirus-covid-19-what-rcr-doing/clinical-information/role-ct-chest/role-ct-screening-0 (accessed 17/05/2020.

24. Jessop ZM, Dobbs TD, Ali SR, et al. Personal Protective Equipment (PPE) for Surgeons during COVID-19 Pandemic: A Systematic Review of Availability, Usage, and Rationing. The British journal of surgery 2020.

25. Improvement N. National Cost Collection for the NHS. 2020. https://improvement.nhs.uk/resources/national-cost-collection/ (accessed 11/05/2020.

26. NICE. Costing statement: Implementing the NICE guideline on Transition between inpatient hospital settings and community or care home settings for adults with social care needs (NG27), 2015.

27. Improvement N. Reference costs 2017/18: highlights, analysis and introduction to the data. 2018.

28. Improvement N. National Tariff Payment System: National prices and prices for emergency care services. In: Improvement N, editor.; 2020.

29. Supplies MM. Medical supplies company. 2020. http://www.medistock.co.uk (accessed 17/05/2020 2020).

30. England N. NHS referral to treatment (RTT) waiting times data March 2020, 2020.

31. Fund TK. The NHS budget and how it has changed. 2020. https://http://www.kingsfund.org.uk/projects/nhs-in-a-nutshell/nhs-budget (accessed 23/05/2020 2020).

32. Leung K, Wu JT, Liu D, Leung GM. First-wave COVID-19 transmissibility and severity in China outside Hubei after control measures, and second-wave scenario planning: a modelling impact assessment. Lancet 2020; 395(10233): 1382–93.

33. Cano-Valderrama O, Morales X, Ferrigni CJ, et al. Reduction in emergency surgery activity during COVID-19 pandemic in three Spanish hospitals. The British journal of surgery 2020.

