## Supplementary material for "Resource requirements for reintroducing elective surgery in England during the COVID-19 pandemic: a modelling study": main tables for manuscript

| **Class of Surgery** | **Mar** | **April** | **May** | **Jun.** | **Jul.** | **Aug.** | **Sep.** | **Oct.** | **Nov.** | **Dec.** | **Jan.** | **Feb.** |
| --- | --- | --- | --- | --- | --- | --- | --- | --- | --- | --- | --- | --- |
| **Procedures carried out each month** | | | | | | | | | | | | |
| **Class 1** | 72 999 (69 230 to 76 769) | 67 776 (63 324 to 72 227) | 70 844 (66 191 to 75 497) | 69 027 (64 493 to 73 561) | 70 502 (65 871 to 75 133) | 67 738 (63 289 to 72 187) | 68 575 (64 071 to 73 079) | 72 048 (67 316 to 76 781) | 71 308 (66 624 to 75 991) | 72 665 (67 892 to 77 438) | 72 593 (67 825 to 77 361) | 66 472 (62 106 to 70 838) |
| **Class 2** | 28 232 (23 362 to 34 218) | 26 566 (21 365 to 33 429) | 27 629 (22 220 to 34 767) | 35 770 (28 767 to 45 010) | 35 854 (28 835 to 45 117) | 33 825 (27 203 to 42 563) | 34 965 (28 120 to 43 998) | 36 731 (29 540 to 46 220) | 36 542 (29 388 to 45 982) | 31 840 (25 607 to 40 066) | 35 094 (28 223 to 44 160) | 33 840 (27 215 to 42 582) |
| **Class 3** | 84 128 (69 109 to 110 982) | 0 (0 to 0) | 0 (0 to 0) | 0 (0 to 0) | 0 (0 to 0) | 0 (0 to 0) | 37 557 (30 940 to 52 842) | 78 907 (65 004 to 111 020) | 11 7752 (97 005 to 165 674) | 136 801 (112 698 to 192 476) | 150 781 (124 216 to 212 146) | 145 392 (119 776 to 204 563) |
| **Class 4** | 58 018 (30 779 to 86 570) | 0 (0 to 0) | 0 (0 to 0) | 0 (0 to 0) | 0 (0 to 0) | 0 (0 to 0) | 0 (0 to 0) | 0 (0 to 0) | 0 (0 to 0) | 28 346 (13 346 to 44 249) | 62 486 (29 419 to 97 542) | 90 380 (42 551 to 141 083) |
| **Total** | 243 377 (192 480 to 308 539) | 94 342 (84 689 to 105 656) | 98 473 (88 411 to 110 264) | 104 797 (93 260 to 118 571) | 106 356 (94 706 to 120 250) | 101 563 (90 492 to 114 750) | 141 097 (123 131 to 169 919) | 18 7686 (161 860 to 234 021) | 225 602 (193 017 to 287 647) | 269 652 (219 543 to 354 229) | 320 954 (249 683 to 431 209) | 336 084 (251 648 to 459 066) |
| **Cumulative deficit of cancelled surgical procedures** | | | | | | | | | | | | |
| **Class 1** | 0 (0 to 0) | 0 (0 to 0) | 0 (0 to 0) | 0 (0 to 0) | 0 (0 to 0) | 0 (0 to 0) | 0 (0 to 0) | 0 (0 to 0) | 0 (0 to 0) | 0 (0 to 0) | 0 (0 to 0) | 0 (0 to 0) |
| **Class 2** | 7 058 (5 840 to 8 555) | 13 699 (11 181 to 16 912) | 20 606 (16 736 to 25 604) | 25 972 (21 051 to 32 356) | 29 557 (23 934 to 36 867) | 31 248 (25 295 to 38 996) | 31 248 (25 295 to 38 996) | 31 248 (25 295 to 38 996) | 31 248 (25 295 to 38 996) | 31 248 (25 294 to 38 996) | 31 248 (25 295 to 38 996) | 31 248 (25 295 to 38 996) |
| **Class 3** | 84 127 (69 109 to 110 982) | 226 800 (186 645 to 311 721) | 375 186 (308 887 to 520 497) | 528 870 (435 494 to 736 727) | 682 917 (562 400 to 953 468) | 828 246 (682 124 to 1 157 943) | 940 917 (774 943 to 1 316 468) | 1 019 824 (839 948 to 1 427 489) | 1 059 074 (872 283 to 1 482 714) | 1 059 074 (872 283 to 1 482 714) | 1 059 074 (872 283 to 1 482 714) | 1 059 074 (872 283 to 1 482 714) |
| **Class 4** | 58 019 (30 779 to 86 569) | 176 272 (86 453 to 271 164) | 299 260 (144 356 to 463 150) | 426 639 (204 326 to 661 990) | 554 319 (264 438 to 861 300) | 674 773 (321 148 to 1 049 330) | 799 287 (379 770 to 1 243 698) | 930 089 (441 352 to 1 447 881) | 1 060 218 (502 617 to 1 651 014) | 1 145 258 (542 653 to 1 783 761) | 1 207 745 (572 072 to 1 881 304) | 1 237 871 (586 256 to 1 928 332) |
| **Total** | 149 204 (105 728 to 206 106) | 416 771 (284 279 to 599 797) | 695 052 (469 979 to 1 009 251) | 981 481 (660 871 to 1 431 073) | 1 266 793 (850 772 to 1 851 635) | 1 534 267 (1 028 567 to 2 246 269) | 1 771 452 (1 180 008 to 2 599 162) | 1 981 161 (1 306 595 to 2 914 366) | 2 150 540 (1 400 195 to 3 172 724) | 2 235 580 (1 440 230 to 3 305 471) | 2 298 067 (1 469 650 to 3 403 014) | 2 328 193 (1 483 834 to 3 450 043) |

**Table 1.** Model for reintroduction of surgical activity between 1^st^ March 2020 and 28^th^ February 2021, assuming: continued class 1 (emergency) activity at pre-pandemic levels; continued class 2 (urgent) activity at 80% of pre-pandemic levels and increasing on 1^st^ June to reach pre-pandemic levels by 31^St^ August; reintroduction of class 3 activity on 1^st^ September and reaching pre-pandemic levels by 31^st^ November; and reintroducing class 4 activity on 30^th^ November and reaching pre-pandemic levels by 28^th^ February 2021. The top panel shows the number of procedures carried out in each month between 1^st^ March 2020 and 28^th^ February 2021. The bottom panel shows the cumulative deficit of surgical activity on the last day of each month. Numbers are presented as predicted time-weighted average with 95% confidence intervals.

| **Item** | **Mar** | **April** | **May** | **Jun.** | **Jul.** | **Aug.** | **Sep.** | **Oct.** | **Nov.** | **Dec.** | **Jan.** | **Feb.** | **Total** |
| --- | --- | --- | --- | --- | --- | --- | --- | --- | --- | --- | --- | --- | --- |
| **Admissions for surgery** | 243 377 (192 480 to 308 539) | 94 342 (84 689 to 105 656) | 98 473 (88 411 to 110 264) | 104 797 (93 260 to 118 571) | 106 356 (94 706 to 120 250) | 101 563 (904 92 to 114 750) | 141 097 (123 131 to 169 919) | 187 686 (161 860 to 234 021) | 225 602 (193 017 to 287 647) | 269 652 (219 543 to 354 229) | 320 954 (249 683 to 431 209) | 336 084 (251 648 to 459 066) | 2 219 341 (1 834 361 to 2 800 729) |
| **Bed days** | 674 874 (575 731 to 792 195) | 454 047 (412 743 to 500 865) | 474 136 (431 057 to 522 959) | 473 547 (429 306 to 524 105) | 487 702 (441 725 to 540 388) | 472 336 (427 424 to 523 916) | 524 948 (471 086 to 595 008) | 592 159 (528 411 to 682 305) | 627 556 (557 139 to 733 557) | 687 956 (590 698 to 825 927) | 771 877 (638 708 to 951 627) | 777 409 (622 654 to 978 739) | 7 018 546 (6 126 681 to 8 171 592) |
| **Day cases** | 107 605 (79 851 to 145 562) | 13 338 (10 727 to 16 783) | 13 871 (11 156 to 17 455) | 15 265 (12 276 to 19 208) | 16 201 (13 029 to 20 387) | 16 133 (12 974 to 20 300) | 46 025 (37 572 to 62 146) | 78 256 (64 107 to 107 364) | 107 608 (88 289 to 148 675) | 134 177 (105 109 to 188 641) | 163 860 (123 369 to 232 850) | 173 404 (126 211 to 248 566) | 885 743 (684 670 to 1 227 936) |
| **Critical care admissions** | 3 549 (3 201 to 3 934) | 2 844 (2 638 to 3 054) | 2 970 (2 759 to 3 193) | 2 911 (2 702 to 3 135) | 2 981 (2 765 to 3 206) | 2 868 (2 661 to 3 089) | 3 009 (2 779 to 3 271) | 3 254 (2 996 to 3 571) | 3 319 (3 045 to 3 670) | 3 537 (3 182 to 3 978) | 3 749 (3 298 to 4 306) | 3 623 (3 118 to 4 229) | 38 614 (35 144 to 42 636) |
| **SARS-CoV 2 PCR test** | 486 754 (384 960 to 617 078) | 188 684 (169 378 to 211 312) | 196 946 (176 822 to 220 528) | 198 862 (177 890 to 223 638) | 205 542 (183 646 to 231 478) | 199 744 (178 262 to 225 242) | 282 194 (246 262 to 339 838) | 375 372 (323 720 to 468 042) | 451 204 (386 034 to 575 294) | 539 304 (439 088 to 708 458) | 641 908 (499 364 to 862 418) | 672 168 (503 296 to 918 132) | 4 438 682 (3 668 722 to 5 601 458) |
| **CT scans** | 23 609 (22 390 to 24 829) | 21 920 (20 480 to 23 360) | 22 912 (21 407 to 24 417) | 22 325 (20 858 to 23 791) | 22 802 (21 304 to 24 299) | 21 908 (20 469 to 23 347) | 22 178 (20 722 to 23 635) | 23 302 (21 771 to 24 832) | 23 062 (21 547 to 24 577) | 23 501 (21 958 to 25 045) | 23 478 (21 936 to 25 020) | 21 498 (20 086 to 22 910) | 272 495 (254 928 to 290 062) |
| **Excess bed days for screening** | 135 772 (112 629 to 162 977) | 81 004 (73 962 to 88 873) | 84 602 (77 255 to 92 809) | 84 166 (76 669 to 92 611) | 86 570 (78 794 to 95 352) | 83 739 (76 157 to 92 321) | 95 072 (85 559 to 107 773) | 109 430 (97 753 to 126 657) | 117 994 (104 728 to 138 972) | 135 475 (114 435 to 165 588) | 157 094 (126 313 to 198 359) | 162 680 (125 437 to 210 500) | 1 333 597 (1 149 690 to 1 572 792) |
| **Excess Cost (low estimate) [€]** | 54 746 570 (44 858 704 to 66 789 186) | 29 103 048 (26 489 800 to 32 053 345) | 30 392 099 (27 666 315 to 33 468 862) | 30 319 814 (27 525 397 to 33 500 255) | 31 214 402 (28 311 986 to 34 526 794) | 30 220 053 (27 386 392 to 33 461 331) | 36 368 613 (32 473 059 to 41 927 260) | 43 756 608 (38 686 430 to 51 854 143) | 48 921 551 (42 913 333 to 59 214 737) | 56 811 493 (47 475 867 to 71 172 800) | 66 267 055 (52 829 664 to 85 442 602) | 68 713 711 (52 636 732 to 90 515 828) | 526 835 017 (449 253 679 to 633 927 143) |
| **Excess cost (high estimate) [€]** | 82 948 422 (67 877 470 to 101 296 839) | 43 897 115 (39 939 812 to 48 370 324) | 45 840 812 (41 713 181 to 50 505 674) | 45 747 560 (41 513 751 to 50 572 273) | 47 102 728 (42 704 526 to 52 128 500) | 45 607 218 (41 312 507 to 50 525 850) | 54 945 177 (49 037 084 to 63 383 763) | 66 148 362 (58 457 243 to 78 441 662) | 73 993 616 (64 878 116 to 89 620 617) | 86 022 467 (71 821 448 to 107 861 022) | 100 469 906 (79 995 877 to 129 666 370) | 104 281 222 (79 762 308 to 137 504 764) | 797 004 605 (679 013 323 to 959 877 658) |

**Table 2.** Resource requirements each month for reintroduction of surgical activity from 1^st^ March 2020 until 28^th^ February 2021. Total number of procedures is presented as a time-weighted average with 95% confidence intervals derived from a linear growth model at procedure group level. Costs are presented in pounds sterling and include only excess costs associated with specific measures required for surgical time. Low estimate: four members of staff in personal protective equipment, bed day cost of £222 (€243.68). High estimate: eight members of staff in personal protective equipment, bed day cost of £346 (€379.79). A complete breakdown of costs is in supplementary table 9. Costs are provided in Euros (€).
