## Supplementary file for "Resource requirements for reintroducing elective surgery in England during the COVID-19 pandemic: a modelling study"

**Appendix**

Unabridged methods	2

Unabridged results	10

Supplementary references	13

Supplementary figures	14

Supplementary tables	15

### Unabridged methods

*Data source*

Aggregated HES data for Admitted Patient Care (APC) between 1^st^ April 2014 and 31^st^ March 2019 (<https://digital.nhs.uk/data-and-information>) were used. HES provide detailed data describing hospital care in England, including the type of surgical procedure categorised by an Office for Population Censuses Surveys Classification of Interventions and Procedures (OPCS) version 4.7 codes. To calculate per month weighting, the monthly activity return from the same time period published by NHS England (<https://www.england.nhs.uk/statistics/statistical-work-areas/hospital-activity/monthly-hospital-activity/mar-data/>) were used. Freely available, anonymised data were used for this study and research ethics and information governance approvals were therefore not required.

*Outcomes*

The primary outcome measures were the deficit of surgical activity attributable to the COVID-19 pandemic response and the estimated volume of surgical procedures upon commencement of non-emergency surgery. The secondary outcomes were the associated resources, defined as: hospital bed days, number of critical care admissions, number of investigations for preoperative COVID-19 screening, the amount of PPE required for operating theatre staff and the additional financial cost.

*Data preparation*

The HES APC includes four categorical variables relating to type of hospital admission: emergency, waiting list, planned and other admission method and these correlate with the method of admission that is allocated to each record during coding. ‘Waiting list’ and ‘planned’ both refer to elective admissions and the distinction between these is based on whether resource availability or clinical need determine the date of admission. For the purposes of this analysis, emergency admissions were the sum of emergency and other admissions; elective admissions were the sum of planned and waiting list. The numbers within HES APC are suppressed when the value is less than a specified figure. By year this varies between five and seven cases and is indicated by an asterisk (*). Suppressed numbers were replaced by the value ‘7’ in all cases, which affected 54 of 342,784 data cells (0·01%). Hyphens ‘-‘ were assumed to be a value of ‘0’. The year 2016 had no ‘0’ values, but 10,230 cells with a hyphen; these were all replaced with ‘0’.

*Modelling analysis*

The statistical analysis plan was prospectively designed and published on our website before data analysis took place.^1^ RStudio, (RStudio Inc., Boston, MA, USA) running on R version 3.6.1 (R Core Team, Vienna, Austria) was used for data analysis.

*Classification of surgical procedures*

Procedures were screened for inclusion by removing codes that were not included in the ‘intermediate’ definition of surgery describe by Abbott et al.^2^ This definition identifies procedures typically performed in an operating theatre under regional or general anaesthesia and constitutes 1047 OPCS 4.7 codes. Since publication in 2017, 35 additional OPCS codes (from version 4.8) have been used in the HES APC data set. These were reviewed by two authors acting independently (AJF, TA) and any discrepancies resolved by discussion. Of these, 27 were added to the intermediate definition of surgery but were excluded from procedure cost analysis as there was no relevant cost code in the 2015 data that was used. Surgical procedures were classified based on lists published by NHS England on 17^th^ March 2020 of operations that should be stopped or continued during the initial pandemic response.^3^ Three investigators (SH, RL, TD) independently reviewed the list of procedure codes. Many procedure classes included repeated OPCS v4.7 codes, for example in some circumstances a joint replacement may need to be carried out within three months of referral and in some circumstances it may be possible to delay this for longer. Thus it was not possible to identify procedures based on independent review of coding alone.

A pre-specified data-driven approach was therefore used to identify emergency, urgent and elective surgical procedures based on waiting times and expert review of procedure coding. Procedure groupings and associated procedure codes are provided in supplementary table 1 and the procedure codes contributing to each class are in supplementary table 2. Reported wait time to classify procedures from HES APC data (2014-2015 to 2018-2019). Both mean and median waiting time are available within the HES APC data set and mean wait time was used for the primary analysis. Surgical procedures were divided into four classes of decreasing urgency: Class 1 – Emergency operations needed within 24 hours and urgent operations needed within 72 hours; Class 2 – Surgery that can be deferred for up to 4 weeks; Class 3 – Surgery that can be delayed for up to 3 months; and Class 4 – Surgery that can be delayed for more than 3 months. Fifty-six procedures had no mean wait time recorded. These were associated with 73 elective admissions over the 5-year period and so excluded from the analysis.

*Number of surgical procedures*

The core analysis was performed at admission level. This was selected as it provides a conservative estimate as each admission had at least one operation meeting a previously described surgical definition as its primary procedure code, and may have had many others. During an admission, an individual may have many finished consultant episodes (FCEs), which are defied by a period spent under the care of a single consultant. The number of day case procedures is expressed in two columns, firstly as the number of episodes associated with a day case procedure and secondly in the ‘elective day case’ column. This is a count of episodes of inpatient admissions that had a zero day length of stay. To determine how many admissions are associated with a day case procedure, the number of day case procedures listed was added to the ‘elective day case’ number, then divided by the number of finished consultant episodes. This provided a proportion of episodes that were day case and we multiplied the number of elective admissions by this to determine how many elective cases were day case. This was not repeated for emergency admissions as it is not possible to have a day case admission for an emergency category record.

*Projected surgical procedure volumes*

The monthly volume of surgical activity that would have been expected through to 31^st^ March 2021 had there not been a pandemic was estimated by calculating the annual change in activity during the five-year period by procedure groupings from 1^st^ April 2014 to 31^st^ March 2019, which was extrapolated using a linear growth assumption. Historical data for this model were from 2014/2015 to 2018/2019. Procedures were grouped according to their anatomical location as in supplementary table 1. Within each procedure group a linear model was developed for each class to determine the regression coefficient and intercept. This model was then used to project future volumes of surgery to 2019/2020 and 2020/2021 for each procedure group and class, including a 95% confidence interval. As it is not possible to have a negative number of procedures, values for procedure projected to be less than zero were truncated to zero. The total number of expected procedures in each year were summed to give an overall procedure volume, subdivided by class.

*Estimating procedural volume by month*

To adjust for monthly variation in elective and emergency admissions, estimates were weighted according to the proportion of elective and emergency admissions reported to NHS England. HES APC data is published on an annual basis, and the available data tables describe activity over a 12-month period between 1^st^ April of one year and 31^st^ March of the subsequent year. Having used the linear growth model to determine the total number of procedures expected in the fiscal year 2020/2021, the total procedure volume was divided it into months by using monthly activity returns (MAR). These are reported to (and available from) NHS England monthly and are admission (not episode) based. Within this, elective total admissions and non-elective total admissions are available on a monthly basis. The count of non-elective includes emergency, maternity and ‘other’ admissions, which aligns with the definition of elective/non-elective implemented during the cleaning of the HES APC data. To calculate the proportion of elective procedures performed by month over the year, he sum of admissions in each month over the 5-year period was divided by the total number of admissions over the 5-year period. This was repeated for non-elective admissions. Values derived from the procedure volume modelling (as outlined above) were then divided using these values. All values were rounded up to the nearest whole number.

*Proportion weighting measures*

To provide a weighted measure of bed days, day case and inpatient admissions the weight that each individual OPCS code contributed to the overall number of admissions was determined by class. For example, for class 2, the count of admissions for each procedure was divided by the 5-year total number of class 2 admissions. This gave each procedure a proportion weighting that it contributed to class 2 overall. To determine the weighted proportion of day case procedures, this number was multiplied by the proportion of patients undergoing day case procedures. As all emergency admissions are overnight admissions, we assumed all class 1 procedures to be inpatient admissions. The proportion of bed days was the proportion cases performed as an inpatient, multiplied by median length of stay. A similar approach was taken to provide a weighting to each class across procedures for each age category. Age is categorised in the data source and these were collated into four groups (0-14, 15-59, 60-74 and 75+ years). The average age was derived for each class of surgery according to the proportion of patients within each age category for each procedure code within a given class.

*Model for reintroduction of surgical activity*

The number of postponed or cancelled surgical procedures from the 1^st^ March 2020 up to 1^st^ June 2020 was estimated according to several assumptions. First, that class 1 (emergency) surgery would continue at the pre-pandemic rate. Second, that class 2 surgery would continue at a reduced rate. We used four scenarios, where 20%, 40%, 60% and 80% of class 2 surgical procedures were assumed to have continued. In the final model we assumed that 80% of class 2 procedures continued. Third, that 50% of class 3 and 4 procedures continued in March and then stopped completely in April and May, reflecting the fact that some hospitals stopped surgery before NHS England advice and some waited until afterwards. The results were presented as the deficit of surgical procedures between 1^st^ March and 31^st^ May 2020 with a 95% confidence interval. Fourth, that widespread reintroduction of surgical activity would start from 1^st^ June 2020 and continue to increase to pre-pandemic levels. For each of the four scenarios of class 2 procedures, we assumed a linear increase in activity over the three months from 1^st^ June to 31^st^ August 2020, with class 3 and 4 surgical procedures remaining cancelled until 31^st^ August. This analysis in an iterative fashion, using the same assumptions, by adding class 3 procedures on 1^st^ September and class 4 procedures on 1^st^ December. Fifth, that pre-pandemic levels of surgical activity would be reached by 28^th^ February 2021 (supplementary figure 1). The estimated number of surgical procedures carried out each month between 1^st^ March 2020 and 28^th^ February 2021 are presented, as is a rolling deficit of surgical activity compared to the expected volume of surgery according to the previous five-year average. Assumptions about a second peak of COVID-19 were not included, and neither were assumptions regarding the impact of reduced operating theatre utilisation due to enhanced infection control procedures. A post-hoc sensitivity analysis was performed assuming that all classes of surgery restarted one the 1^st^ June 2020 and increased linearly over a 6-month period.

Restart of activity was from the 31^st^ May to 31^st^ August for class 2 procedures, from 31^st^ August to 30^st^ November for class 3 procedures and from 30^th^ November to the 28^th^ February to class 4 procedures. Linear growth was modelled from the baseline of zero activity for classes 3 & 4 and 80% of normal activity for class 2. This was calculated such that 1/4 of normal activity occurred in the first build up month, 2/4 in the second, 3/4 in the third and full activity resumed by the end of the month after the build up period.

*Hospital admissions*

The total number of bed-days was calculated, weighted by procedure frequency within class, by multiplying the median length of stay by the number of inpatient admissions. The proportion of patients in each class of surgery that were likely to require a postoperative critical care bed was estimated using a conservative estimate of 1% of those undergoing inpatient elective surgery, and 4% of those undergoing emergency surgery, based on previously published data.^4-6^

*Preoperative screening tests*

The burden of preoperative screening for COVID-19 was estimated according to three scenarios. First, all patients would have two preoperative COVID-19 Polymerase Chain Reaction (PCR) tests performed on an outpatient basis. Second, all patients would have two preoperative SARS-COV-2 PCR tests, one on an outpatient basis and one on an inpatient basis, requiring an additional day of inpatient stay for preoperative isolation. Third, all patients would have two preoperative SARS-COV-2 PCR tests, one on an outpatient basis and one on an inpatient basis. In addition, patients undergoing class 1 (emergency) thoracic, cardiac or abdominal procedures would also have a computed tomography scan of the chest, in line with guidance from the Royal College of Radiologists, to identify radiological signs of COVID-19 in patients with an increased likelihood of postoperative critical care unit admission.^7^^,^ ^8^

*Personal protective equipment*

It is advised that full personal protective equipment will be required in operating theatres where patients receive mechanical ventilation or undergo aerosol-generating procedures. The amount of personal protective equipment required per month was estimated according to two scenarios.^9^ First, that eight members of staff (two surgeons, two anaesthetists, one operating department practitioner and three scrub staff) would be present for every surgical procedure, and second, that four members of staff (one surgeon, one anaesthetist, one operating department practitioner and one scrub staff) for each procedure, each requiring an FFP3 mask, a fluid repellent gown, two pairs of gloves and a face shield or visor.

*Estimated financial cost*

The estimated financial cost of reintroducing surgical activity was divided into three areas: the cost of the surgical procedure; the cost of preoperative COVID-19 screening arrangements; and, the cost of PPE required. The total cost of reintroducing surgical activity was estimated from 1^st^ June 2020 and the total deficit of surgical procedures on 1^st^ March 2021 was estimated by combining these. Costs were calculated in British Pounds (£) and are presented in Euros (€) based on the average exchange rate reported by OANDA on the 1^st^ July 2020 (£1 = €1.09766). The total costs of surgical procedures was calculated by matching the OPCS v4.7 code with the Health Resource Group coding using a previously published method, and summing the associated procedure cost according to the national schedule of NHS costs in 2015 as described in our prior work.^2^^,^ ^10^ The costs of screening tests were calculated according to £19 (€20.86) per SARS-COV-2 PCR and £69 (€75.74) per CT scan and a range of between £222 (€243.68) and £346 (€379.79) per additional bed day.^11-13^ The costs of PPE was calculated as £2.90 (€3.18) per FFP3 mask, £14.90 (€16.36) per 100 gloves, £3 (€3.29) per fluid resistant gown and £2.90 (€3.18) per piece of eye protection.^13^^,^ ^14^

*Role of the funding source*

There is no funding to report for this study. AF and TA had full access to all the data in the study and all authors had final responsibility for the decision to submit for publication.

### Unabridged results

Some 1073 OPCS codes for Class 1 – 4 surgical procedures, representing a total of 22 513 872 surgical admissions between 1^st^ April 2014 and 31^st^ March 2019 (Figure 1). Over the five-year period, the monthly median number of procedures was 382 768 (Interquartile range [IQR]: 22 890). The weighted five-year median number of procedures per month for each class of surgery is listed in supplementary table 3. If growth in the number of surgical procedures had continued according to a pre-pandemic growth trajectory, the estimated number of procedures for the year 1^st^ March 2020 to 28^th^ February 2021 would be 4 547 534 (95%CI: 3 318 195 to 6 250 771) (supplementary table 4). The proportion of patients in each age grouping in a typical month is presented by class in supplementary table 5. Patients aged over 60 years account for 32.7% of class 1, 43.9% of class 2, 53.7% of class 3 and 45.6% of class 4 activity.

*Surgical procedure volume*

If all class 1 (emergency) procedures continued, with 80% of class 2 procedures in March and 50% of class 3 and 4 procedures in March, the estimated number of cancelled operations by 31^st^ May 2020 in comparison with observed pre-pandemic levels of activity, which is termed ‘deficit’, would be 749 247 (95%CI: 513 564 to 1 077 448) surgical procedures (supplementary table 6). If both class 1 (emergency) and class 2 (urgent) procedures continued at pre-pandemic levels, the deficit of surgical activity on 31^st^ May 2020 would be 674 446 (95%CI: 453 243 to 983 648) procedures. However, it is assumed that lower than usual class 2 (urgent) activity would have occurred between March and May 2020. Therefore the widespread reintroduction of class 2 surgical activity from 31^st^ May 2020, was modelled for four scenarios of incrementally increasing activity (20% - 80%) that would reach pre-pandemic levels by 31^st^ August 2020 (supplementary table 7). If all class 1 surgery and 80% of class 2 surgery continued during the pandemic response, increasing to pre-pandemic levels of activity by 31^st^ August 2020, the total deficit of surgery would be 1 534 267 (95%CI: 1 028,567 to 2 246 269) procedures (Table 1). If class 3 surgical activity is reintroduced from 31^st^ August 2020 and reached pre-pandemic level of activity by 30^th^ November 2020, the total deficit of surgical activity on 30^th^ November 2020 would be 2 150 540 (95%CI: 1 400 195 to 3 172 724) procedures. If class 4 surgical activity is reintroduced from 31^st^ November 2020 and reached a pre-pandemic level of activity by 28^th^ February 2021, the deficit of surgical activity on that date would be 2 328 193 (95%CI: 1 483 834 to 3 450 043) procedures (table 1, figures 2-3). In a post-hoc sensitivity analysis assuming classes 2,3 and 4 restarted on 1^st^ June and took six months to get to normal capacity, the number of cancelled surgical procedures was estimated to be 1 551 560 (95%CI: 1 041 537 to 2 269 486) (supplementary table 11).

*Hospital admissions*

The total bed days associated with the deficit of surgical activity on 31^st^ May 2020, assuming that 80% of class 2 activity was undertaken are 973 006 (95%CI: 623 700 to 1 423 014) days (supplementary table 8). The number of bed days required for the continuation of class 1 surgical activity and reintroduction of class 2 surgical activity from 31^st^ May, class 3 activity from the 31^st^ August and class 4 activity from 30^th^ November is provided in table 2. If widespread reintroduction of surgery happens according to this schedule, the estimated total bed days associated with the cumulative deficit of surgical activity up to 28^th^ February 2021 is 3 337 706 days (95%CI: 1 997 510 to 4 995 117). The estimated number of patients requiring day case procedures is 1 451 295 (95%CI: 973 616 to 2 129 276). The estimated total number of critical care admissions associated with the deficit of surgical activity on 31^st^ May 2020 is 2 474 (95%CI: 1 536 to 3 650) (supplementary table 8). If widespread reintroduction of surgical activity occurs from 31^st^ May, the estimated total number of critical care admissions associated with the cumulative deficit of surgery up to 28th February 2021 will be 8 769 admissions (95%CI: 5 103 to 13 207).

*Preoperative screening tests*

The estimated total number of SARS-CoV-2 PCR tests and additional bed days for preoperative isolation associated with the deficit of surgical activity on 31^st^ May 2020 are: 1 390 104 (95%CI: 939 958 to 2 018 502) tests, and 247 321 (95%CI: 153 633 to 365 080) bed days respectively. The estimated total number of SARS-CoV-2 PCR tests and additional bed days for preoperative isolation associated with the reintroduction of surgical activity from 1^st^ June 2020 is provided in table 2.

*Personal protective equipment*

The estimated total amount of personal protective equipment (PPE) associated with the deficit of surgical activity on 31^st^ May 2020 is 11 120 832 (95% CI: 7 519 664 to 16 148 016) items, assuming four persons wear PPE, and 22 241 664 (95% CI: 15 039 328 to 32 296 032) items assuming eight persons wear PPE. The estimated total cost of PPE associated with reintroducing surgical activity from 31^st^ May is provided in supplementary table 8. The estimated total amount of PPE associated with the cumulative deficit of surgical activity by 28th February 2021 is between 37 251 088 (95%CI: 23 741 344 to 55 200 672) items, assuming four persons per theatre and 74 502 176 items (95%CI: 47 482 688 to 110 401 344) assuming eight persons per theatre (supplementary table 8).

*Financial cost*

The procedure cost for the deficit of surgical activity on 31^st^ May 2020 is €1.5B (95%CI: €923.0M to €2.2B) (supplementary table 10). The cost of personal protective equipment and preoperative screening for COVID-19 associated with recommencing surgical activity from 31^st^ May is given in table 2 and supplementary table 9. The financial costs associated with reintroducing elective surgery on 31^st^ June 2020 until 28^th^ February 2021, including the cost of the surgical procedure, personal protective equipment and screening, is between €5.4B (95%CI: €4.4B to €6.7B) and €5.7B (95%CI: €4.7B to €7.0B) (table 2 and supplementary table 9). If elective surgery recommences on 1^st^ June 2020 in a stepwise fashion, the total cost of cancelled or delayed operations on 28^th^ February 2021, including the cost of the surgical procedure, PPE and screening tests, will be between €5.8B (95%CI: €3.3B to €8.6B) and €5.9B (95%CI: €3.5B to €8.9B) (supplementary table 10).

### Supplementary references

1.	Fowler AJ, Dobbs TD, Abbott TEF. Estimated surgical requirements in England after COVID-19: a modelling study using hospital episode statistics. 2020. https://<http://www.qmul.ac.uk/ccpmg/sops--saps/statistical-analysis-plans-saps/09/05/2020)>.

2.	Abbott TEF, Fowler AJ, Dobbs TD, Harrison EM, Gillies MA, Pearse RM. Frequency of surgical treatment and related hospital procedures in the UK: a national ecological study using hospital episode statistics. *British journal of anaesthesia* 2017; **119**(2): 249-57.

3.	NHS England. Clinical guide to surgical prioritsation during the coronavirus pandemic, 2020.

4.	International Surgical Outcomes Study Group. Global patient outcomes after elective surgery: prospective cohort study in 27 low-, middle- and high-income countries. *British journal of anaesthesia* 2016; **117**(5): 601-9.

5.	Pearse RM, Moreno RP, Bauer P, et al. Mortality after surgery in Europe: a 7 day cohort study. *Lancet* 2012; **380**(9847): 1059-65.

6.	Intensive Care National Audit and Research Centre. Summary Statistics. 2019. https://<http://www.icnarc.org/Our-Audit/Audits/Cmp/Reports/Summary-Statistics> (accessed 20/05/2020 2020).

7.	Lima DS, Ribeiro MAF, Jr., Gallo G, Di Saverio S. Role of chest CT in patients with acute abdomen during the COVID-19 era. *The British journal of surgery* 2020.

8.	The Royal College of Radiologists. Statement on use of CT chest to screen for COVID-19 in pre-operative patients. 14/05/2020 2020. https://<http://www.rcr.ac.uk/college/coronavirus-covid-19-what-rcr-doing/clinical-information/role-ct-chest/role-ct-screening-0> (accessed 17/05/2020.

9.	Jessop ZM, Dobbs TD, Ali SR, et al. Personal Protective Equipment (PPE) for Surgeons during COVID-19 Pandemic: A Systematic Review of Availability, Usage, and Rationing. *The British journal of surgery* 2020.

10.	NHS Improvement. National Cost Collection for the NHS. 2020. https://improvement.nhs.uk/resources/national-cost-collection/ (accessed 11/05/2020.

11.	National Institute for health and Care Excellence. Costing statement: Implementing the NICE guideline on Transition between inpatient hospital settings and community or care home settings for adults with social care needs (NG27), 2015.

12.	NHS Improvement. Reference costs 2017/18: highlights, analysis and introduction to the data. 2018.

13.	NHS Improvement. National Tariff Payment System: National prices and prices for emergency care services. In: Improvement N, editor.; 2020.

14.	Medical supplies company. 2020. <http://www.medistock.co.uk> (accessed 17/05/2020 2020).

### Supplementary figures

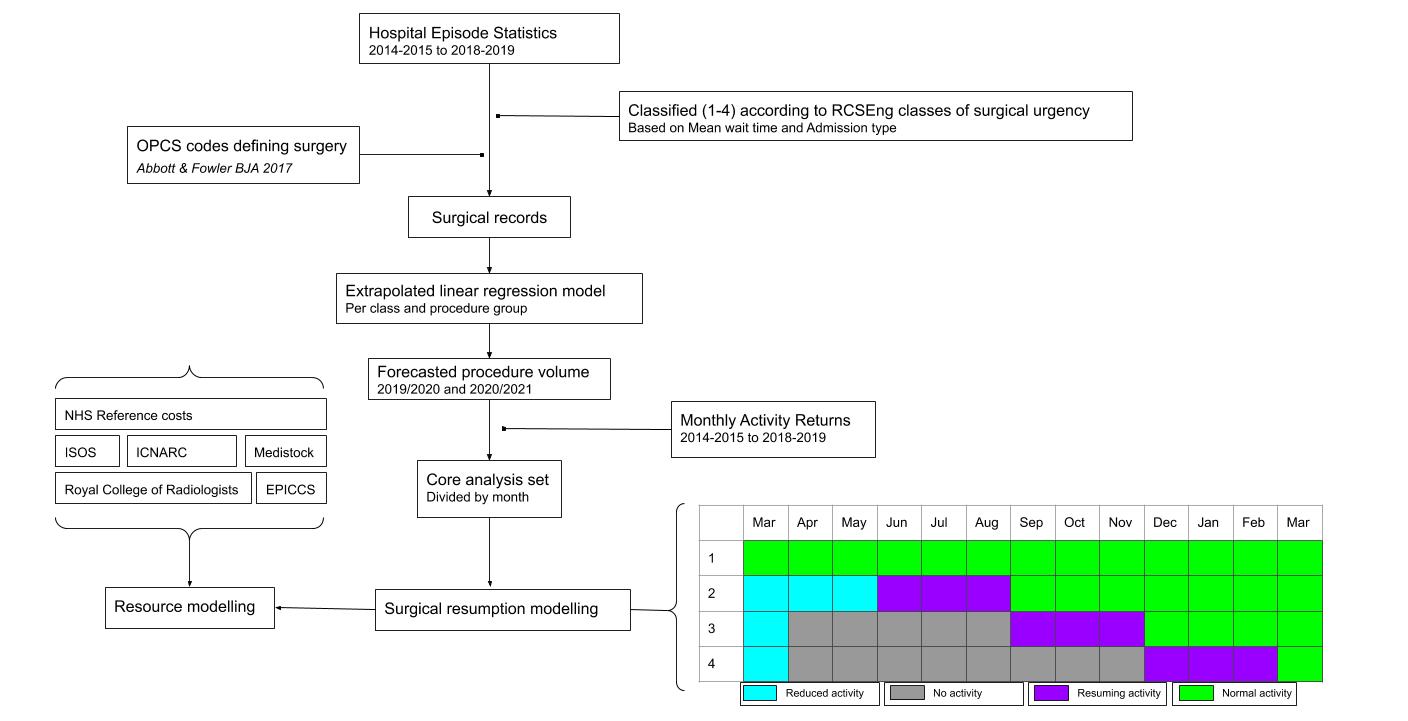

**Supplementary figure 1. Detailed flow diagram of analysis methodology and assumptions.**

### Supplementary tables

| **Grouping** | **OPCS codes** |
| --- | --- |
| Neurosurgery | A06, A82, A16, A20, A22, A47, A05, A41, A11, A08, A04, A42, A45, A40, A01, A18, A12, A02, A10, A07, A76, A03, A43, A17, A38, A14, A84, A44, A79, A51, A13, A28, A39, A29, A78, A81, A75, A73, A59, A32, A60, A48, A49, A36, A68, A09, A57, A69, A26, A33, A62, A70, A65, A64, A67, A61, A63, A30, A24, A66, A25, A31, A34, A27, A77 |
| Ocular | C23, C16, C01, C17, C20, C26, C10, C06, C74, C09, C02, C51, C34, C84, C15, C73, C66, C69, C82, C75, C22, C47, C14, C64, C59, C08, C79, C43, C77, C40, C71, C46, C11, C88, C62, C86, C24, C89, C49, C19, C54, C44, C37, C29, C52, C81, C05, C33, C61, C25, C39, C18, C13, C35, C12, C80, C60, C27, C65, C45, C32, C31, C85, C57, C03, C67, C55, C72, C53, C41, C83 |
| Nasal | E64, E65, E66, E05, E01, E15, E17, E11, E10, E16, E12, E09, E13, E08, E14, E04, E03, E02, E07 |
| Upper GI | G26, G12, G20, G57, G71, G11, G52, G10, G35, G58, G61, G69, G36, G74, G53, G63, G41, G78, G49, G67, G76, G60, G59, G17, G14, G34, G73, G07, G27, G06, G72, G08, G31, G05, G33, T37, T15, G51, G82, G01, G13, G38, G28, G03, G21, G70, G50, T16, G02, G68, G23, G29, T17, G75, G48, G04, G32, G40, G25, G24, G09, G30 |
| Lower GI | H32, H37, T32, T30, H13, H30, H29, H11, H17, T36, H08, H05, H19, H14, H06, T45, T41, H15, H09, H07, H16, H33, H10, T34, T48, T38, T39, T22, T28, T31, H47, H44, H04, T33, T23, H62, X14, H36, H12, T51, H46, T29, T43, T25, T27, H42, H03, H41, T26, H40, H34, T21, H54, H53, H35, T97, H58, T24, T42, T20, T98, H66, H49, H50, H01, H56, H52, H02, H55, H51, T19, H48, H60, H59, H57 |
| Cardiac | K72, K73, K74, K71, K56, K55, K77, K68, K53, K32, K54, K69, K15, K01, K02, K44, K30, K33, K17, K40, K07, K43, K36, K24, K38, K41, K35, K23, K27, K67, K18, K25, L02, K47, K19, K46, K26, K14, K31, K12, K29, K52, K60, K34, K11, K06, K75, K45, K22, K48, K65, K20, K64, K08, K04, K13, K66, K09, K28, K59, K16, K76, K37, K10, K57, K62, K05, K42, K78 |
| Joint | O32, W47, W48, W46, W57, W39, W49, W80, W64, W51, W37, W67, W85, W81, W92, W95, W65, W42, W93, W50, W91, W38, W94, W98, W96, W86, T62, W69, W40, W41, W45, W43, W97, W55, W44, W61, W60, W63, W58, W62, W88, W87, W56, W78, W71, W89, W83, W84, W77, W59, W82, W79, W70, W01 |
| Female UGU | R06, Q57, Q50, Q56, Q51, Q47, Q20, Q22, Q07, Q52, Q44, Q24, Q09, Q43, Q36, Q27, Q23, Q54, Q08, Q10, Q17, Q49, Q16, Q01, Q02, Q41, Q35, Q11, Q25, Q05, Q38, Q31, Q30, Q39, Q32, Q37, Q29, Q45, Q28, Q34, Q26, Q19 |
| Skin | S63, T61, S40, S37, S57, S41, S17, S54, S56, S35, S55, S47, S24, S33, S42, S18, S10, S25, S11, S36, S30, S26, S27, S06, S62, S31, S60, S70, S04, S02, S68, S01, S05, S48, S23, S66, S64, T59, S49, T60, S39, S03, S22, S20, S19, S21, S38, S28, S34, T94 |
| Skull and spine | V02, V12, V18, V51, V03, V05, V04, V47, V24, V06, V46, V43, V31, V44, V45, V60, V14, V40, V23, V37, V20, V22, V07, V52, V41, V19, V01, V29, V21, V38, V30, V08, V25, V42, V26, V35, V48, V17, V39, V15, V27, V34, V11, V54, V13, V16, V36, V67, V33, V49, V10, V09, V28, V68, V32, V62, V56, V66, V63, V58, V61, V57 |
| Other | X16, X04, X17, X55, T88, W99, T87, T86, T89, W34, T92, T96, T85, T91, X46, X53, X03 |
| Breast | B41, B37, B27, B33, B35, B28, B30, B34, B29, B36, B31, B39, B38, B40 |
| Thoracic | E67, T13, T12, E42, T03, T10, E44, E52, E50, E48, T08, T14, T09, E47, E53, T11, E59, T05, E61, E63, E57, E41, E62, E54, E43, T07, E55, T01, E40, E46, E39, T02 |
| Female LGU | M57, P28, P30, P29, P18, P17, P09, P31, P11, P06, P22, P05, P13, P24, P20, P23, P25, P03, P19, P15, P07, P21, P01, P14, P32 |
| Major Vessel | L18, L48, L46, L20, L25, L45, L22, L42, L50, L16, L47, L05, L79, L08, L53, L12, L06, L38, L13, L19, L21, L23, L07, L41, L30, L80, L49, L26, L04, L01, L77, L10, L28, L69, L27, L52, L39, L51, L37, L43, L31, L09, L54, L29 |
| HPB | J04, J33, J24, J06, J05, J15, J21, J07, J13, J25, J11, J23, J16, J31, J19, J65, J72, J57, J60, J30, J52, J69, J49, J12, J01, J55, J26, J58, J10, J56, J61, J32, J35, J34, J68, J36, J29, J02, J70, J37, J20, J73, J03, J54, J27, J59, J08, J18, J28, J63, J77, J62 |
| Vascular | L62, L71, X09, O15, L81, L99, L58, L91, L56, X07, L90, L96, O01, L97, O02, L65, O03, X10, L60, X12, L59, L76, L63, X11, L70, L93, L66, L94, O04, L57, L74, L82, L83, L68, L75, X08, L67, L03, L85, L86, L88, L87, L84, L98, O20, L73, L89 |
| Cerebrovascular | L34, L33, O05, L35 |
| Urological | M06, M15, M60, M33, M48, M13, M37, M26, M38, M16, M41, M08, M22, M34, M19, M68, M35, M67, M02, M25, M29, M44, M39, M83, M17, M42, M21, M23, M01, M04, M18, M62, M55, M27, M49, M20, M65, M64, M86, M10, M32, M79, M03, M76, M28, M71, M81, M66, M72, M75, X15, M58, M09, M61, M05, M70, M43, M73, M56, M53, M52, M36, M54, M51 |
| Endocrine | B25, B12, B09, B06, B04, B01, B22, B18, B14, B16, B08, B10, B23, B02, B17, B20 |
| Bone | W24, W19, W29, W05, W25, W33, W22, X20, W18, W20, X19, W26, W53, W30, W23, W52, W28, W21, W17, W09, W04, W08, X25, W54, X27, W02, W06, W31, X21, W13, X22, W03, W16, W15, X24, W14, W27, W12, X23, W68, W32, W10, W07, W11, X05, X02, X01 |
| Pharynx | E19, E33, E29, E28, E31, E35, E27, E23, E30, E24, E38, E21, E34, E20, F34 |
| Muscle | T55, T83, T77, T53, T76, T54, T65, T57, W73, T67, T70, T72, T74, T64, T52, T79, T68, T56, T69, W76, W75, W74, T80, T71, W72, T50 |
| Male GU | N05, N24, N26, N03, N01, N06, N32, N30, N13, N27, N11, N29, N28, N15, N09, N17, N08, N07, N19, N10, N34, N20, N18, N22 |
| Oral | F58, F48, F22, F39, F46, F24, F40, F53, F01, F42, F38, F32, F30, F06, F23, F04, F44, F36, F05, F28, F45, F26, F02, F11, F09, F29, F03, F18, F51, F50, F52 |
| Organ donation | X45 |
| Orthopaedics | O23, O22, O17, O18, O25, O21, O07, O27, O29, O26, O19, O06, O24, O10, O08, O09 |
| Ear | D04, D06, D01, D08, D02, D28, D10, D19, D24, D12, D15, D20, D13, D03, D14, D16, D17, D26, D23, D22 |
| Obstetrics | R30, R34, R18, R12, R17, R29, R28, R07, R08, R01, R10, R05, R04, R02 |

**Supplementary table 1. Office of Population Censuses and Surveys (OPCS) codes defining surgery, with associated procedure groupings.** Selected according to a previously published method.^2^ Thirty-five additional codes were identified that were reported in admitted patient care data since the prior definition was constructed. These were screened by two authors acting independently and 27 were selected for inclusion. UGU; upper genitourinary tract, HPB; hepatopancreatobiliary, GU; genitourinary tract, GI; gastrointestinal, LGU; lower genitourinary tract.

| OPCS | class1 | class2 | class3 | class4 |
| --- | --- | --- | --- | --- |
| A01 | 5 | 0 | 4 | 1 |
| A02 | 5 | 4 | 1 | 0 |
| A03 | 4 | 0 | 2 | 3 |
| A04 | 5 | 5 | 0 | 0 |
| A05 | 5 | 0 | 5 | 0 |
| A06 | 5 | 0 | 5 | 0 |
| A07 | 5 | 0 | 2 | 3 |
| A08 | 5 | 5 | 0 | 0 |
| A09 | 5 | 0 | 5 | 0 |
| A10 | 5 | 5 | 0 | 0 |
| A11 | 5 | 0 | 5 | 0 |
| A12 | 5 | 0 | 5 | 0 |
| A13 | 5 | 0 | 5 | 0 |
| A14 | 5 | 0 | 5 | 0 |
| A16 | 5 | 1 | 4 | 0 |
| A17 | 5 | 0 | 5 | 0 |
| A18 | 5 | 2 | 3 | 0 |
| A20 | 5 | 0 | 5 | 0 |
| A22 | 5 | 2 | 3 | 0 |
| A24 | 3 | 0 | 0 | 5 |
| A25 | 2 | 0 | 1 | 4 |
| A26 | 5 | 0 | 5 | 0 |
| A27 | 3 | 1 | 4 | 0 |
| A28 | 4 | 0 | 5 | 0 |
| A29 | 5 | 0 | 4 | 1 |
| A30 | 5 | 0 | 4 | 1 |
| A31 | 2 | 0 | 5 | 0 |
| A32 | 5 | 0 | 0 | 5 |
| A33 | 5 | 0 | 4 | 1 |
| A34 | 5 | 0 | 4 | 1 |
| A36 | 5 | 0 | 2 | 3 |
| A38 | 5 | 0 | 5 | 0 |
| A39 | 5 | 0 | 3 | 2 |
| A40 | 5 | 1 | 3 | 1 |
| A41 | 5 | 5 | 0 | 0 |
| A42 | 5 | 0 | 5 | 0 |
| A43 | 5 | 0 | 5 | 0 |
| A44 | 5 | 0 | 5 | 0 |
| A45 | 5 | 1 | 4 | 0 |
| A47 | 3 | 5 | 0 | 0 |
| A48 | 5 | 0 | 4 | 1 |
| A49 | 5 | 0 | 3 | 2 |
| A51 | 5 | 0 | 1 | 4 |
| A57 | 5 | 0 | 2 | 3 |
| A59 | 5 | 0 | 3 | 2 |
| A60 | 5 | 0 | 1 | 4 |
| A61 | 5 | 0 | 3 | 2 |
| A62 | 5 | 5 | 0 | 0 |
| A63 | 5 | 0 | 5 | 0 |
| A64 | 5 | 5 | 0 | 0 |
| A65 | 5 | 0 | 5 | 0 |
| A66 | 2 | 0 | 1 | 4 |
| A67 | 5 | 0 | 4 | 1 |
| A68 | 5 | 0 | 3 | 2 |
| A69 | 5 | 0 | 3 | 2 |
| A70 | 5 | 0 | 2 | 3 |
| A73 | 5 | 0 | 1 | 4 |
| A75 | 5 | 0 | 4 | 1 |
| A76 | 5 | 0 | 3 | 2 |
| A77 | 2 | 0 | 5 | 0 |
| A78 | 3 | 0 | 1 | 4 |
| A79 | 2 | 0 | 2 | 3 |
| A81 | 5 | 0 | 1 | 4 |
| A84 | 5 | 0 | 5 | 0 |
| B01 | 5 | 0 | 5 | 0 |
| B02 | 1 | 1 | 1 | 1 |
| B04 | 5 | 0 | 5 | 0 |
| B06 | 5 | 3 | 2 | 0 |
| B08 | 5 | 0 | 5 | 0 |
| B09 | 0 | 0 | 4 | 1 |
| B10 | 5 | 0 | 2 | 3 |
| B12 | 5 | 1 | 4 | 0 |
| B14 | 5 | 0 | 2 | 3 |
| B16 | 5 | 0 | 2 | 3 |
| B17 | 0 | 1 | 1 | 0 |
| B18 | 5 | 0 | 5 | 0 |
| B20 | 5 | 3 | 2 | 0 |
| B22 | 5 | 0 | 5 | 0 |
| B23 | 0 | 3 | 2 | 0 |
| B25 | 5 | 5 | 0 | 0 |
| B27 | 5 | 0 | 5 | 0 |
| B28 | 5 | 5 | 0 | 0 |
| B29 | 5 | 0 | 0 | 5 |
| B30 | 5 | 0 | 4 | 1 |
| B31 | 5 | 0 | 0 | 5 |
| B33 | 5 | 0 | 5 | 0 |
| B34 | 5 | 0 | 5 | 0 |
| B35 | 5 | 0 | 5 | 0 |
| B36 | 5 | 0 | 0 | 5 |
| B37 | 5 | 0 | 0 | 5 |
| B38 | 1 | 1 | 0 | 4 |
| B39 | 5 | 0 | 0 | 5 |
| B40 | 2 | 1 | 4 | 0 |
| C01 | 5 | 0 | 5 | 0 |
| C02 | 5 | 0 | 4 | 1 |
| C03 | 5 | 0 | 3 | 2 |
| C05 | 5 | 0 | 2 | 3 |
| C06 | 5 | 0 | 5 | 0 |
| C08 | 5 | 3 | 2 | 0 |
| C09 | 4 | 0 | 2 | 3 |
| C10 | 5 | 0 | 5 | 0 |
| C11 | 5 | 0 | 5 | 0 |
| C12 | 5 | 0 | 5 | 0 |
| C13 | 4 | 0 | 0 | 5 |
| C14 | 5 | 0 | 5 | 0 |
| C15 | 5 | 0 | 5 | 0 |
| C16 | 5 | 0 | 5 | 0 |
| C17 | 5 | 0 | 5 | 0 |
| C18 | 5 | 0 | 0 | 5 |
| C19 | 5 | 0 | 5 | 0 |
| C20 | 5 | 0 | 5 | 0 |
| C22 | 5 | 0 | 5 | 0 |
| C23 | 5 | 0 | 2 | 3 |
| C24 | 5 | 0 | 5 | 0 |
| C25 | 5 | 0 | 0 | 5 |
| C26 | 5 | 0 | 5 | 0 |
| C27 | 5 | 0 | 4 | 1 |
| C29 | 5 | 0 | 5 | 0 |
| C31 | 5 | 0 | 0 | 5 |
| C32 | 5 | 0 | 0 | 5 |
| C33 | 4 | 0 | 0 | 5 |
| C34 | 3 | 0 | 1 | 4 |
| C35 | 5 | 0 | 0 | 5 |
| C37 | 5 | 0 | 2 | 3 |
| C39 | 5 | 0 | 5 | 0 |
| C40 | 5 | 0 | 5 | 0 |
| C41 | 5 | 0 | 4 | 1 |
| C43 | 5 | 0 | 5 | 0 |
| C44 | 5 | 0 | 0 | 5 |
| C45 | 5 | 0 | 5 | 0 |
| C46 | 5 | 0 | 0 | 5 |
| C47 | 5 | 0 | 5 | 0 |
| C49 | 5 | 0 | 5 | 0 |
| C51 | 5 | 0 | 5 | 0 |
| C52 | 5 | 0 | 5 | 0 |
| C53 | 2 | 0 | 5 | 0 |
| C54 | 5 | 5 | 0 | 0 |
| C55 | 5 | 3 | 2 | 0 |
| C57 | 5 | 3 | 2 | 0 |
| C59 | 5 | 1 | 4 | 0 |
| C60 | 5 | 0 | 5 | 0 |
| C61 | 5 | 0 | 5 | 0 |
| C62 | 5 | 0 | 5 | 0 |
| C64 | 5 | 0 | 5 | 0 |
| C65 | 5 | 0 | 5 | 0 |
| C66 | 5 | 0 | 5 | 0 |
| C67 | 4 | 1 | 4 | 0 |
| C69 | 5 | 4 | 1 | 0 |
| C71 | 5 | 0 | 5 | 0 |
| C72 | 5 | 0 | 2 | 3 |
| C73 | 5 | 0 | 5 | 0 |
| C74 | 5 | 0 | 5 | 0 |
| C75 | 5 | 0 | 5 | 0 |
| C77 | 5 | 0 | 5 | 0 |
| C79 | 5 | 0 | 5 | 0 |
| C80 | 5 | 0 | 5 | 0 |
| C81 | 5 | 5 | 0 | 0 |
| C82 | 5 | 0 | 5 | 0 |
| C83 | 0 | 1 | 1 | 1 |
| C84 | 5 | 1 | 4 | 0 |
| C85 | 5 | 5 | 0 | 0 |
| C86 | 5 | 0 | 5 | 0 |
| C88 | 4 | 3 | 1 | 1 |
| C89 | 5 | 0 | 3 | 2 |
| D01 | 5 | 0 | 5 | 0 |
| D02 | 5 | 0 | 5 | 0 |
| D03 | 5 | 0 | 0 | 5 |
| D04 | 5 | 5 | 0 | 0 |
| D06 | 5 | 0 | 5 | 0 |
| D08 | 5 | 0 | 5 | 0 |
| D10 | 5 | 0 | 0 | 5 |
| D12 | 5 | 0 | 0 | 5 |
| D13 | 5 | 0 | 0 | 5 |
| D14 | 5 | 0 | 0 | 5 |
| D15 | 5 | 0 | 4 | 1 |
| D16 | 4 | 0 | 0 | 5 |
| D17 | 5 | 0 | 0 | 5 |
| D19 | 5 | 0 | 5 | 0 |
| D20 | 5 | 0 | 5 | 0 |
| D22 | 3 | 0 | 1 | 4 |
| D23 | 5 | 0 | 5 | 0 |
| D24 | 5 | 0 | 4 | 1 |
| D26 | 5 | 0 | 3 | 2 |
| D28 | 5 | 0 | 5 | 0 |
| E01 | 4 | 2 | 3 | 0 |
| E02 | 5 | 0 | 0 | 5 |
| E03 | 5 | 0 | 2 | 3 |
| E04 | 5 | 0 | 2 | 3 |
| E05 | 5 | 0 | 5 | 0 |
| E07 | 5 | 0 | 0 | 5 |
| E08 | 5 | 0 | 4 | 1 |
| E09 | 5 | 0 | 5 | 0 |
| E10 | 5 | 0 | 5 | 0 |
| E11 | 5 | 0 | 5 | 0 |
| E12 | 5 | 0 | 5 | 0 |
| E13 | 5 | 0 | 2 | 3 |
| E14 | 5 | 0 | 2 | 3 |
| E15 | 5 | 0 | 2 | 3 |
| E16 | 5 | 0 | 3 | 2 |
| E17 | 5 | 0 | 4 | 1 |
| E19 | 5 | 5 | 0 | 0 |
| E20 | 5 | 0 | 0 | 5 |
| E21 | 5 | 0 | 0 | 5 |
| E23 | 5 | 0 | 5 | 0 |
| E24 | 5 | 0 | 5 | 0 |
| E27 | 5 | 0 | 5 | 0 |
| E28 | 5 | 0 | 2 | 3 |
| E29 | 5 | 5 | 0 | 0 |
| E30 | 5 | 0 | 5 | 0 |
| E31 | 5 | 0 | 5 | 0 |
| E33 | 5 | 0 | 5 | 0 |
| E34 | 5 | 0 | 5 | 0 |
| E35 | 5 | 0 | 5 | 0 |
| E38 | 5 | 0 | 5 | 0 |
| E39 | 5 | 0 | 5 | 0 |
| E40 | 5 | 0 | 5 | 0 |
| E41 | 5 | 0 | 5 | 0 |
| E42 | 5 | 0 | 5 | 0 |
| E43 | 5 | 0 | 3 | 2 |
| E44 | 2 | 1 | 2 | 0 |
| E46 | 5 | 1 | 4 | 0 |
| E47 | 5 | 3 | 2 | 0 |
| E48 | 5 | 1 | 4 | 0 |
| E50 | 5 | 4 | 1 | 0 |
| E52 | 5 | 2 | 2 | 1 |
| E53 | 5 | 1 | 0 | 4 |
| E54 | 5 | 5 | 0 | 0 |
| E55 | 5 | 0 | 5 | 0 |
| E57 | 5 | 1 | 4 | 0 |
| E59 | 5 | 5 | 0 | 0 |
| E61 | 5 | 2 | 3 | 0 |
| E62 | 5 | 0 | 4 | 1 |
| E63 | 5 | 5 | 0 | 0 |
| E64 | 5 | 0 | 4 | 1 |
| F01 | 5 | 0 | 5 | 0 |
| F02 | 5 | 0 | 5 | 0 |
| F03 | 5 | 0 | 0 | 5 |
| F04 | 5 | 0 | 4 | 1 |
| F05 | 5 | 0 | 5 | 0 |
| F06 | 5 | 0 | 5 | 0 |
| F09 | 5 | 0 | 5 | 0 |
| F11 | 5 | 0 | 1 | 4 |
| F18 | 5 | 0 | 5 | 0 |
| F22 | 5 | 3 | 2 | 0 |
| F23 | 5 | 0 | 5 | 0 |
| F24 | 5 | 5 | 0 | 0 |
| F26 | 5 | 0 | 5 | 0 |
| F28 | 5 | 0 | 5 | 0 |
| F29 | 5 | 0 | 0 | 5 |
| F30 | 5 | 0 | 0 | 5 |
| F32 | 5 | 0 | 5 | 0 |
| F34 | 5 | 0 | 2 | 3 |
| F36 | 5 | 0 | 5 | 0 |
| F38 | 5 | 0 | 5 | 0 |
| F39 | 5 | 0 | 3 | 2 |
| F40 | 5 | 0 | 5 | 0 |
| F42 | 5 | 0 | 5 | 0 |
| F44 | 5 | 0 | 5 | 0 |
| F45 | 5 | 0 | 5 | 0 |
| F46 | 5 | 0 | 5 | 0 |
| F48 | 5 | 0 | 5 | 0 |
| F50 | 0 | 0 | 0 | 5 |
| F51 | 5 | 0 | 4 | 1 |
| F52 | 2 | 0 | 1 | 4 |
| F53 | 5 | 0 | 3 | 2 |
| F58 | 5 | 0 | 4 | 1 |
| G01 | 5 | 3 | 2 | 0 |
| G02 | 5 | 3 | 2 | 0 |
| G03 | 5 | 5 | 0 | 0 |
| G04 | 4 | 0 | 5 | 0 |
| G05 | 3 | 0 | 4 | 1 |
| G06 | 5 | 1 | 3 | 1 |
| G07 | 5 | 0 | 5 | 0 |
| G08 | 5 | 0 | 5 | 0 |
| G09 | 5 | 0 | 2 | 3 |
| G10 | 5 | 0 | 4 | 1 |
| G11 | 5 | 2 | 3 | 0 |
| G13 | 5 | 3 | 2 | 0 |
| G14 | 5 | 0 | 5 | 0 |
| G17 | 5 | 0 | 5 | 0 |
| G21 | 5 | 0 | 5 | 0 |
| G23 | 5 | 0 | 0 | 5 |
| G24 | 5 | 0 | 0 | 5 |
| G25 | 5 | 0 | 0 | 5 |
| G26 | 5 | 0 | 0 | 0 |
| G27 | 5 | 5 | 0 | 0 |
| G28 | 5 | 0 | 2 | 3 |
| G29 | 5 | 0 | 5 | 0 |
| G30 | 5 | 0 | 5 | 0 |
| G31 | 5 | 0 | 2 | 3 |
| G32 | 5 | 0 | 2 | 3 |
| G33 | 5 | 0 | 2 | 3 |
| G34 | 5 | 0 | 5 | 0 |
| G35 | 5 | 1 | 4 | 0 |
| G36 | 5 | 0 | 3 | 2 |
| G38 | 5 | 0 | 4 | 1 |
| G40 | 5 | 0 | 5 | 0 |
| G41 | 5 | 4 | 1 | 0 |
| G48 | 5 | 0 | 5 | 0 |
| G49 | 5 | 1 | 4 | 0 |
| G50 | 5 | 0 | 5 | 0 |
| G51 | 5 | 1 | 4 | 0 |
| G52 | 5 | 1 | 2 | 1 |
| G53 | 5 | 0 | 5 | 0 |
| G57 | 5 | 2 | 3 | 0 |
| G58 | 5 | 0 | 5 | 0 |
| G59 | 5 | 2 | 3 | 0 |
| G60 | 5 | 1 | 4 | 0 |
| G61 | 5 | 0 | 3 | 2 |
| G63 | 5 | 1 | 4 | 0 |
| G67 | 5 | 2 | 3 | 0 |
| G68 | 5 | 1 | 0 | 0 |
| G69 | 5 | 0 | 5 | 0 |
| G70 | 5 | 0 | 5 | 0 |
| G71 | 5 | 0 | 5 | 0 |
| G72 | 5 | 0 | 0 | 5 |
| G73 | 5 | 0 | 5 | 0 |
| G74 | 5 | 0 | 5 | 0 |
| G75 | 5 | 0 | 3 | 2 |
| G76 | 5 | 0 | 5 | 0 |
| G78 | 5 | 0 | 4 | 1 |
| G82 | 5 | 1 | 4 | 0 |
| H01 | 5 | 0 | 5 | 0 |
| H02 | 5 | 0 | 5 | 0 |
| H03 | 5 | 0 | 0 | 5 |
| H04 | 5 | 0 | 5 | 0 |
| H05 | 5 | 0 | 5 | 0 |
| H06 | 5 | 5 | 0 | 0 |
| H07 | 5 | 4 | 1 | 0 |
| H08 | 5 | 0 | 5 | 0 |
| H09 | 5 | 1 | 4 | 0 |
| H10 | 5 | 0 | 5 | 0 |
| H11 | 5 | 0 | 5 | 0 |
| H12 | 5 | 0 | 5 | 0 |
| H13 | 5 | 2 | 3 | 0 |
| H14 | 5 | 0 | 5 | 0 |
| H15 | 5 | 0 | 5 | 0 |
| H16 | 5 | 2 | 3 | 0 |
| H17 | 5 | 0 | 4 | 1 |
| H19 | 5 | 0 | 5 | 0 |
| H29 | 5 | 0 | 5 | 0 |
| H30 | 5 | 0 | 4 | 1 |
| H32 | 5 | 0 | 1 | 4 |
| H33 | 5 | 0 | 5 | 0 |
| H34 | 5 | 0 | 2 | 3 |
| H35 | 5 | 0 | 0 | 5 |
| H36 | 5 | 0 | 1 | 4 |
| H40 | 5 | 0 | 5 | 0 |
| H41 | 5 | 0 | 5 | 0 |
| H42 | 5 | 0 | 1 | 4 |
| H44 | 5 | 0 | 5 | 0 |
| H46 | 5 | 0 | 5 | 0 |
| H47 | 4 | 0 | 2 | 3 |
| H48 | 5 | 0 | 5 | 0 |
| H49 | 5 | 0 | 5 | 0 |
| H50 | 5 | 0 | 1 | 4 |
| H51 | 5 | 0 | 4 | 1 |
| H52 | 5 | 0 | 5 | 0 |
| H53 | 5 | 0 | 4 | 1 |
| H54 | 5 | 0 | 5 | 0 |
| H55 | 5 | 0 | 5 | 0 |
| H56 | 5 | 0 | 5 | 0 |
| H57 | 4 | 0 | 1 | 4 |
| H58 | 5 | 1 | 4 | 0 |
| H59 | 5 | 0 | 4 | 1 |
| H60 | 5 | 0 | 5 | 0 |
| H62 | 5 | 0 | 5 | 0 |
| H66 | 5 | 0 | 3 | 2 |
| J01 | 5 | 0 | 1 | 4 |
| J02 | 5 | 0 | 5 | 0 |
| J03 | 5 | 0 | 5 | 0 |
| J04 | 5 | 2 | 3 | 0 |
| J05 | 5 | 2 | 3 | 0 |
| J06 | 5 | 4 | 1 | 0 |
| J07 | 4 | 1 | 4 | 0 |
| J08 | 5 | 0 | 5 | 0 |
| J10 | 5 | 5 | 0 | 0 |
| J11 | 5 | 4 | 1 | 0 |
| J12 | 5 | 5 | 0 | 0 |
| J13 | 5 | 5 | 0 | 0 |
| J15 | 5 | 3 | 2 | 0 |
| J16 | 5 | 3 | 2 | 0 |
| J18 | 5 | 0 | 4 | 1 |
| J19 | 5 | 0 | 2 | 3 |
| J20 | 5 | 0 | 5 | 0 |
| J21 | 5 | 0 | 4 | 1 |
| J23 | 5 | 0 | 5 | 0 |
| J24 | 5 | 5 | 0 | 0 |
| J25 | 5 | 5 | 0 | 0 |
| J26 | 5 | 2 | 3 | 0 |
| J27 | 5 | 0 | 5 | 0 |
| J28 | 5 | 0 | 5 | 0 |
| J29 | 5 | 0 | 5 | 0 |
| J30 | 5 | 0 | 5 | 0 |
| J31 | 5 | 0 | 4 | 1 |
| J32 | 5 | 1 | 3 | 1 |
| J33 | 5 | 0 | 5 | 0 |
| J34 | 5 | 0 | 5 | 0 |
| J35 | 5 | 5 | 0 | 0 |
| J36 | 4 | 1 | 4 | 0 |
| J37 | 5 | 0 | 5 | 0 |
| J49 | 4 | 1 | 3 | 1 |
| J52 | 5 | 3 | 2 | 0 |
| J54 | 5 | 1 | 0 | 0 |
| J55 | 5 | 0 | 5 | 0 |
| J56 | 5 | 5 | 0 | 0 |
| J57 | 5 | 0 | 5 | 0 |
| J58 | 5 | 0 | 5 | 0 |
| J59 | 5 | 0 | 5 | 0 |
| J60 | 5 | 3 | 2 | 0 |
| J61 | 5 | 5 | 0 | 0 |
| J62 | 0 | 0 | 1 | 0 |
| J63 | 1 | 3 | 1 | 0 |
| J65 | 5 | 5 | 0 | 0 |
| J68 | 4 | 4 | 1 | 0 |
| J69 | 5 | 0 | 5 | 0 |
| J70 | 5 | 0 | 5 | 0 |
| J72 | 5 | 5 | 0 | 0 |
| J73 | 4 | 4 | 1 | 0 |
| K01 | 5 | 1 | 0 | 2 |
| K02 | 5 | 0 | 3 | 2 |
| K04 | 5 | 0 | 5 | 0 |
| K05 | 2 | 1 | 2 | 2 |
| K06 | 5 | 0 | 2 | 3 |
| K07 | 5 | 1 | 3 | 1 |
| K08 | 5 | 0 | 4 | 1 |
| K09 | 5 | 0 | 5 | 0 |
| K10 | 5 | 0 | 4 | 1 |
| K11 | 5 | 0 | 5 | 0 |
| K12 | 5 | 0 | 2 | 3 |
| K13 | 5 | 0 | 1 | 4 |
| K14 | 5 | 0 | 3 | 2 |
| K15 | 4 | 2 | 1 | 1 |
| K16 | 5 | 0 | 5 | 0 |
| K17 | 5 | 0 | 0 | 5 |
| K18 | 5 | 0 | 0 | 5 |
| K19 | 5 | 0 | 1 | 4 |
| K20 | 5 | 0 | 3 | 2 |
| K22 | 5 | 0 | 5 | 0 |
| K23 | 5 | 1 | 4 | 0 |
| K24 | 5 | 0 | 2 | 3 |
| K25 | 5 | 0 | 1 | 4 |
| K26 | 5 | 0 | 5 | 0 |
| K27 | 5 | 0 | 0 | 5 |
| K28 | 5 | 0 | 0 | 5 |
| K29 | 3 | 0 | 2 | 3 |
| K30 | 5 | 0 | 4 | 1 |
| K31 | 5 | 0 | 5 | 0 |
| K32 | 0 | 0 | 1 | 0 |
| K33 | 5 | 0 | 0 | 5 |
| K34 | 5 | 0 | 2 | 3 |
| K35 | 5 | 0 | 5 | 0 |
| K36 | 1 | 2 | 1 | 2 |
| K37 | 4 | 0 | 5 | 0 |
| K38 | 5 | 0 | 5 | 0 |
| K40 | 5 | 0 | 5 | 0 |
| K41 | 5 | 0 | 5 | 0 |
| K42 | 3 | 0 | 2 | 1 |
| K43 | 0 | 1 | 2 | 0 |
| K44 | 5 | 0 | 4 | 1 |
| K45 | 5 | 0 | 5 | 0 |
| K46 | 3 | 0 | 2 | 2 |
| K47 | 5 | 0 | 3 | 2 |
| K48 | 5 | 0 | 5 | 0 |
| K52 | 5 | 0 | 3 | 2 |
| K53 | 5 | 1 | 2 | 2 |
| K54 | 5 | 3 | 2 | 0 |
| K55 | 5 | 0 | 5 | 0 |
| K56 | 5 | 0 | 5 | 0 |
| K57 | 5 | 0 | 0 | 5 |
| K59 | 5 | 0 | 5 | 0 |
| K60 | 5 | 0 | 5 | 0 |
| K62 | 5 | 0 | 0 | 5 |
| K64 | 5 | 0 | 1 | 4 |
| K65 | 5 | 0 | 5 | 0 |
| K66 | 5 | 1 | 4 | 0 |
| K67 | 5 | 0 | 5 | 0 |
| K68 | 5 | 1 | 3 | 1 |
| K69 | 5 | 3 | 2 | 0 |
| K71 | 5 | 2 | 3 | 0 |
| K72 | 5 | 0 | 5 | 0 |
| K75 | 5 | 0 | 5 | 0 |
| K76 | 5 | 0 | 2 | 3 |
| K77 | 5 | 0 | 4 | 1 |
| K78 | 2 | 1 | 3 | 0 |
| L01 | 5 | 0 | 5 | 0 |
| L02 | 5 | 0 | 5 | 0 |
| L03 | 5 | 0 | 3 | 2 |
| L04 | 5 | 1 | 0 | 4 |
| L05 | 3 | 0 | 2 | 3 |
| L06 | 4 | 0 | 3 | 2 |
| L07 | 5 | 0 | 4 | 1 |
| L08 | 5 | 0 | 4 | 1 |
| L09 | 5 | 0 | 5 | 0 |
| L10 | 5 | 0 | 4 | 1 |
| L12 | 5 | 1 | 3 | 1 |
| L13 | 5 | 0 | 2 | 3 |
| L16 | 5 | 0 | 5 | 0 |
| L18 | 5 | 1 | 4 | 0 |
| L19 | 5 | 0 | 5 | 0 |
| L20 | 5 | 2 | 3 | 0 |
| L21 | 5 | 0 | 5 | 0 |
| L22 | 5 | 0 | 5 | 0 |
| L23 | 5 | 0 | 5 | 0 |
| L25 | 5 | 0 | 5 | 0 |
| L26 | 5 | 0 | 4 | 1 |
| L27 | 5 | 0 | 5 | 0 |
| L28 | 5 | 0 | 5 | 0 |
| L29 | 5 | 5 | 0 | 0 |
| L30 | 5 | 0 | 5 | 0 |
| L31 | 5 | 1 | 4 | 0 |
| L33 | 5 | 0 | 5 | 0 |
| L34 | 5 | 0 | 5 | 0 |
| L35 | 5 | 0 | 5 | 0 |
| L37 | 5 | 0 | 5 | 0 |
| L38 | 5 | 1 | 4 | 0 |
| L39 | 5 | 0 | 5 | 0 |
| L41 | 4 | 0 | 4 | 1 |
| L42 | 5 | 2 | 3 | 0 |
| L43 | 5 | 0 | 5 | 0 |
| L45 | 5 | 2 | 3 | 0 |
| L46 | 5 | 1 | 4 | 0 |
| L47 | 5 | 0 | 5 | 0 |
| L48 | 5 | 2 | 1 | 1 |
| L49 | 5 | 0 | 5 | 0 |
| L50 | 5 | 1 | 4 | 0 |
| L51 | 5 | 0 | 5 | 0 |
| L52 | 5 | 0 | 5 | 0 |
| L53 | 5 | 0 | 5 | 0 |
| L54 | 5 | 0 | 5 | 0 |
| L56 | 5 | 3 | 2 | 0 |
| L57 | 5 | 0 | 5 | 0 |
| L58 | 5 | 2 | 3 | 0 |
| L59 | 5 | 0 | 5 | 0 |
| L60 | 5 | 0 | 5 | 0 |
| L62 | 5 | 0 | 5 | 0 |
| L63 | 5 | 0 | 5 | 0 |
| L65 | 5 | 2 | 3 | 0 |
| L66 | 5 | 0 | 5 | 0 |
| L67 | 5 | 5 | 0 | 0 |
| L68 | 5 | 3 | 2 | 0 |
| L69 | 5 | 0 | 3 | 2 |
| L70 | 5 | 0 | 2 | 3 |
| L71 | 5 | 0 | 5 | 0 |
| L73 | 2 | 2 | 2 | 1 |
| L74 | 5 | 0 | 5 | 0 |
| L75 | 5 | 0 | 5 | 0 |
| L76 | 1 | 2 | 2 | 0 |
| L77 | 4 | 1 | 2 | 2 |
| L79 | 5 | 1 | 4 | 0 |
| L80 | 4 | 0 | 4 | 1 |
| L81 | 5 | 1 | 4 | 0 |
| L82 | 3 | 0 | 4 | 1 |
| L83 | 4 | 0 | 2 | 3 |
| L84 | 5 | 0 | 2 | 3 |
| L85 | 5 | 0 | 1 | 4 |
| L86 | 5 | 0 | 2 | 3 |
| L87 | 5 | 0 | 1 | 4 |
| L88 | 5 | 0 | 1 | 4 |
| L89 | 0 | 2 | 0 | 0 |
| L90 | 5 | 0 | 5 | 0 |
| L91 | 5 | 5 | 0 | 0 |
| L93 | 5 | 0 | 5 | 0 |
| L94 | 5 | 4 | 1 | 0 |
| L96 | 5 | 4 | 1 | 0 |
| L97 | 5 | 0 | 3 | 2 |
| L98 | 5 | 0 | 1 | 4 |
| L99 | 5 | 5 | 0 | 0 |
| M01 | 5 | 0 | 5 | 0 |
| M02 | 5 | 0 | 5 | 0 |
| M03 | 5 | 0 | 5 | 0 |
| M04 | 4 | 0 | 3 | 2 |
| M05 | 5 | 0 | 3 | 2 |
| M06 | 5 | 0 | 5 | 0 |
| M08 | 5 | 2 | 3 | 0 |
| M09 | 5 | 0 | 5 | 0 |
| M10 | 5 | 0 | 5 | 0 |
| M13 | 5 | 4 | 1 | 0 |
| M15 | 5 | 5 | 0 | 0 |
| M16 | 5 | 0 | 5 | 0 |
| M17 | 5 | 5 | 0 | 0 |
| M18 | 4 | 0 | 5 | 0 |
| M19 | 5 | 0 | 2 | 3 |
| M20 | 5 | 0 | 2 | 3 |
| M21 | 5 | 0 | 5 | 0 |
| M22 | 5 | 0 | 3 | 2 |
| M23 | 5 | 0 | 5 | 0 |
| M25 | 5 | 0 | 4 | 1 |
| M26 | 5 | 0 | 5 | 0 |
| M27 | 5 | 0 | 5 | 0 |
| M28 | 5 | 0 | 5 | 0 |
| M29 | 5 | 0 | 5 | 0 |
| M32 | 5 | 0 | 4 | 1 |
| M33 | 5 | 2 | 3 | 0 |
| M34 | 5 | 0 | 5 | 0 |
| M35 | 5 | 0 | 5 | 0 |
| M36 | 0 | 0 | 0 | 5 |
| M37 | 5 | 0 | 4 | 1 |
| M38 | 5 | 0 | 3 | 2 |
| M39 | 5 | 0 | 2 | 3 |
| M41 | 5 | 0 | 4 | 1 |
| M42 | 5 | 0 | 5 | 0 |
| M43 | 5 | 0 | 0 | 5 |
| M44 | 5 | 0 | 4 | 1 |
| M48 | 5 | 0 | 2 | 3 |
| M49 | 5 | 0 | 5 | 0 |
| M51 | 2 | 0 | 0 | 5 |
| M52 | 4 | 0 | 0 | 5 |
| M53 | 5 | 0 | 0 | 5 |
| M54 | 4 | 0 | 2 | 3 |
| M55 | 4 | 0 | 1 | 4 |
| M56 | 5 | 0 | 0 | 5 |
| M58 | 5 | 0 | 5 | 0 |
| M60 | 5 | 0 | 3 | 2 |
| M61 | 5 | 0 | 5 | 0 |
| M62 | 3 | 0 | 4 | 1 |
| M64 | 5 | 0 | 0 | 5 |
| M65 | 5 | 0 | 1 | 4 |
| M66 | 5 | 0 | 2 | 3 |
| M67 | 5 | 0 | 5 | 0 |
| M68 | 2 | 0 | 3 | 2 |
| M70 | 5 | 0 | 5 | 0 |
| M71 | 4 | 0 | 5 | 0 |
| M72 | 5 | 0 | 0 | 5 |
| M73 | 5 | 0 | 0 | 5 |
| M75 | 5 | 0 | 5 | 0 |
| M76 | 5 | 0 | 5 | 0 |
| M79 | 5 | 0 | 5 | 0 |
| M81 | 5 | 0 | 5 | 0 |
| M83 | 5 | 5 | 0 | 0 |
| M86 | 5 | 0 | 5 | 0 |
| N01 | 5 | 0 | 5 | 0 |
| N03 | 5 | 0 | 5 | 0 |
| N05 | 5 | 0 | 5 | 0 |
| N06 | 5 | 0 | 5 | 0 |
| N07 | 5 | 0 | 3 | 2 |
| N08 | 5 | 0 | 0 | 5 |
| N09 | 5 | 0 | 0 | 5 |
| N10 | 5 | 0 | 0 | 5 |
| N11 | 5 | 0 | 2 | 3 |
| N13 | 5 | 0 | 4 | 1 |
| N15 | 5 | 0 | 2 | 3 |
| N17 | 5 | 0 | 4 | 1 |
| N18 | 5 | 0 | 3 | 2 |
| N19 | 5 | 0 | 5 | 0 |
| N20 | 5 | 0 | 5 | 0 |
| N22 | 3 | 0 | 5 | 0 |
| N24 | 5 | 0 | 5 | 0 |
| N26 | 5 | 5 | 0 | 0 |
| N27 | 5 | 0 | 5 | 0 |
| N28 | 5 | 0 | 0 | 5 |
| N29 | 5 | 0 | 0 | 5 |
| N30 | 5 | 0 | 3 | 2 |
| N32 | 5 | 0 | 5 | 0 |
| N34 | 5 | 0 | 2 | 3 |
| O01 | 5 | 0 | 5 | 0 |
| O02 | 5 | 0 | 5 | 0 |
| O03 | 5 | 0 | 5 | 0 |
| O04 | 5 | 0 | 5 | 0 |
| O05 | 5 | 0 | 5 | 0 |
| O06 | 5 | 0 | 0 | 5 |
| O07 | 5 | 0 | 0 | 5 |
| O08 | 3 | 0 | 0 | 5 |
| O09 | 3 | 0 | 1 | 4 |
| O10 | 5 | 0 | 3 | 2 |
| O15 | 5 | 3 | 0 | 0 |
| O17 | 5 | 4 | 1 | 0 |
| O18 | 5 | 0 | 0 | 5 |
| O19 | 5 | 0 | 4 | 1 |
| O21 | 5 | 0 | 0 | 5 |
| O22 | 5 | 0 | 1 | 4 |
| O23 | 5 | 0 | 4 | 1 |
| O24 | 5 | 0 | 4 | 1 |
| O25 | 5 | 2 | 3 | 0 |
| O26 | 5 | 0 | 5 | 0 |
| O27 | 5 | 0 | 0 | 5 |
| O29 | 5 | 0 | 2 | 3 |
| P01 | 5 | 0 | 5 | 0 |
| P03 | 5 | 0 | 5 | 0 |
| P05 | 5 | 0 | 5 | 0 |
| P06 | 5 | 0 | 5 | 0 |
| P07 | 5 | 0 | 5 | 0 |
| P09 | 5 | 0 | 5 | 0 |
| P11 | 5 | 0 | 5 | 0 |
| P13 | 5 | 0 | 5 | 0 |
| P14 | 2 | 0 | 2 | 3 |
| P15 | 5 | 0 | 5 | 0 |
| P17 | 5 | 0 | 5 | 0 |
| P18 | 5 | 0 | 1 | 4 |
| P19 | 5 | 0 | 5 | 0 |
| P20 | 5 | 0 | 5 | 0 |
| P21 | 5 | 0 | 0 | 5 |
| P22 | 5 | 0 | 1 | 4 |
| P23 | 5 | 0 | 0 | 5 |
| P24 | 5 | 0 | 0 | 5 |
| P25 | 5 | 0 | 2 | 3 |
| P29 | 5 | 0 | 5 | 0 |
| P31 | 5 | 0 | 4 | 1 |
| P32 | 2 | 0 | 0 | 5 |
| Q01 | 5 | 0 | 5 | 0 |
| Q02 | 5 | 0 | 5 | 0 |
| Q05 | 5 | 0 | 5 | 0 |
| Q07 | 5 | 0 | 5 | 0 |
| Q08 | 5 | 0 | 4 | 1 |
| Q09 | 5 | 0 | 1 | 4 |
| Q10 | 5 | 0 | 5 | 0 |
| Q11 | 5 | 5 | 0 | 0 |
| Q16 | 5 | 0 | 5 | 0 |
| Q17 | 5 | 0 | 5 | 0 |
| Q19 | 0 | 1 | 3 | 1 |
| Q20 | 5 | 0 | 5 | 0 |
| Q22 | 5 | 0 | 5 | 0 |
| Q23 | 5 | 0 | 5 | 0 |
| Q24 | 5 | 0 | 5 | 0 |
| Q25 | 5 | 0 | 5 | 0 |
| Q26 | 0 | 0 | 2 | 2 |
| Q27 | 5 | 0 | 4 | 1 |
| Q28 | 5 | 0 | 5 | 0 |
| Q29 | 1 | 0 | 5 | 0 |
| Q30 | 5 | 0 | 1 | 4 |
| Q31 | 5 | 0 | 4 | 1 |
| Q32 | 5 | 0 | 5 | 0 |
| Q34 | 5 | 0 | 3 | 2 |
| Q35 | 5 | 0 | 4 | 1 |
| Q36 | 4 | 0 | 4 | 1 |
| Q37 | 3 | 0 | 4 | 1 |
| Q38 | 5 | 0 | 4 | 1 |
| Q39 | 5 | 1 | 4 | 0 |
| Q41 | 5 | 0 | 5 | 0 |
| Q43 | 5 | 0 | 5 | 0 |
| Q44 | 5 | 0 | 4 | 1 |
| Q45 | 5 | 0 | 5 | 0 |
| Q47 | 5 | 0 | 5 | 0 |
| Q49 | 5 | 0 | 4 | 1 |
| Q50 | 5 | 0 | 5 | 0 |
| Q51 | 5 | 2 | 3 | 0 |
| Q52 | 5 | 0 | 4 | 1 |
| Q54 | 5 | 0 | 1 | 4 |
| Q56 | 5 | 0 | 2 | 3 |
| R01 | 5 | 1 | 0 | 0 |
| R02 | 4 | 0 | 0 | 0 |
| R04 | 5 | 3 | 0 | 0 |
| R05 | 5 | 4 | 0 | 0 |
| R06 | 5 | 5 | 0 | 0 |
| R07 | 5 | 4 | 0 | 0 |
| R08 | 5 | 2 | 0 | 0 |
| R10 | 5 | 5 | 0 | 0 |
| R12 | 5 | 5 | 0 | 0 |
| R17 | 5 | 1 | 4 | 0 |
| R18 | 5 | 4 | 1 | 0 |
| R28 | 5 | 5 | 0 | 0 |
| R29 | 5 | 5 | 0 | 0 |
| R30 | 5 | 5 | 0 | 0 |
| R34 | 5 | 5 | 0 | 0 |
| S01 | 5 | 0 | 0 | 5 |
| S02 | 5 | 0 | 0 | 5 |
| S03 | 4 | 0 | 0 | 5 |
| S04 | 5 | 0 | 4 | 1 |
| S05 | 5 | 0 | 2 | 3 |
| S06 | 5 | 0 | 5 | 0 |
| S10 | 5 | 0 | 5 | 0 |
| S11 | 5 | 0 | 5 | 0 |
| S17 | 5 | 0 | 4 | 1 |
| S18 | 5 | 0 | 2 | 3 |
| S19 | 5 | 1 | 2 | 2 |
| S20 | 5 | 0 | 2 | 3 |
| S21 | 5 | 3 | 2 | 0 |
| S22 | 5 | 2 | 3 | 0 |
| S23 | 5 | 0 | 0 | 5 |
| S24 | 5 | 0 | 5 | 0 |
| S25 | 5 | 0 | 5 | 0 |
| S26 | 5 | 0 | 5 | 0 |
| S27 | 5 | 0 | 5 | 0 |
| S28 | 1 | 2 | 2 | 0 |
| S30 | 5 | 0 | 5 | 0 |
| S31 | 5 | 0 | 1 | 4 |
| S33 | 2 | 0 | 0 | 5 |
| S34 | 0 | 0 | 1 | 0 |
| S35 | 5 | 3 | 2 | 0 |
| S36 | 5 | 0 | 5 | 0 |
| S37 | 5 | 0 | 5 | 0 |
| S38 | 0 | 0 | 4 | 1 |
| S39 | 5 | 0 | 1 | 4 |
| S40 | 5 | 4 | 1 | 0 |
| S41 | 5 | 5 | 0 | 0 |
| S42 | 5 | 5 | 0 | 0 |
| S47 | 5 | 5 | 0 | 0 |
| S48 | 5 | 0 | 0 | 5 |
| S49 | 5 | 0 | 5 | 0 |
| S54 | 5 | 4 | 1 | 0 |
| S55 | 5 | 4 | 1 | 0 |
| S56 | 5 | 4 | 1 | 0 |
| S57 | 5 | 3 | 2 | 0 |
| S60 | 5 | 0 | 0 | 5 |
| S62 | 5 | 0 | 0 | 5 |
| S64 | 5 | 0 | 5 | 0 |
| S66 | 5 | 5 | 0 | 0 |
| S68 | 5 | 0 | 5 | 0 |
| S70 | 5 | 0 | 5 | 0 |
| T01 | 5 | 0 | 5 | 0 |
| T02 | 5 | 0 | 0 | 5 |
| T03 | 5 | 0 | 5 | 0 |
| T05 | 5 | 0 | 5 | 0 |
| T07 | 5 | 5 | 0 | 0 |
| T08 | 5 | 4 | 1 | 0 |
| T09 | 5 | 5 | 0 | 0 |
| T10 | 5 | 5 | 0 | 0 |
| T11 | 5 | 5 | 0 | 0 |
| T12 | 5 | 5 | 0 | 0 |
| T13 | 5 | 4 | 1 | 0 |
| T14 | 5 | 5 | 0 | 0 |
| T15 | 5 | 1 | 2 | 1 |
| T16 | 5 | 0 | 5 | 0 |
| T17 | 5 | 1 | 4 | 0 |
| T19 | 5 | 0 | 5 | 0 |
| T20 | 5 | 0 | 5 | 0 |
| T21 | 5 | 0 | 3 | 2 |
| T22 | 5 | 0 | 5 | 0 |
| T23 | 5 | 0 | 4 | 1 |
| T24 | 5 | 0 | 4 | 1 |
| T25 | 5 | 0 | 0 | 5 |
| T26 | 5 | 0 | 0 | 5 |
| T27 | 5 | 0 | 3 | 2 |
| T28 | 5 | 0 | 0 | 5 |
| T29 | 5 | 0 | 5 | 0 |
| T30 | 5 | 0 | 5 | 0 |
| T31 | 5 | 0 | 5 | 0 |
| T33 | 5 | 0 | 5 | 0 |
| T34 | 5 | 0 | 5 | 0 |
| T36 | 5 | 5 | 0 | 0 |
| T37 | 5 | 1 | 4 | 0 |
| T38 | 5 | 0 | 5 | 0 |
| T39 | 5 | 3 | 2 | 0 |
| T41 | 5 | 0 | 5 | 0 |
| T42 | 5 | 0 | 4 | 1 |
| T43 | 5 | 0 | 5 | 0 |
| T45 | 5 | 5 | 0 | 0 |
| T48 | 5 | 4 | 1 | 0 |
| T50 | 4 | 0 | 2 | 3 |
| T51 | 5 | 0 | 2 | 3 |
| T52 | 5 | 0 | 2 | 3 |
| T53 | 5 | 0 | 5 | 0 |
| T54 | 5 | 0 | 4 | 1 |
| T55 | 5 | 0 | 1 | 4 |
| T56 | 4 | 0 | 0 | 5 |
| T57 | 5 | 0 | 5 | 0 |
| T59 | 5 | 0 | 5 | 0 |
| T60 | 2 | 0 | 4 | 1 |
| T61 | 5 | 0 | 5 | 0 |
| T62 | 5 | 0 | 5 | 0 |
| T64 | 5 | 0 | 0 | 5 |
| T65 | 5 | 0 | 5 | 0 |
| T67 | 5 | 2 | 3 | 0 |
| T68 | 5 | 0 | 5 | 0 |
| T69 | 5 | 0 | 2 | 3 |
| T70 | 5 | 0 | 0 | 5 |
| T71 | 5 | 0 | 5 | 0 |
| T72 | 5 | 0 | 5 | 0 |
| T74 | 5 | 0 | 5 | 0 |
| T76 | 5 | 0 | 0 | 5 |
| T77 | 5 | 0 | 5 | 0 |
| T79 | 5 | 0 | 5 | 0 |
| T80 | 5 | 0 | 0 | 5 |
| T83 | 5 | 0 | 5 | 0 |
| T85 | 5 | 5 | 0 | 0 |
| T86 | 5 | 5 | 0 | 0 |
| T87 | 5 | 5 | 0 | 0 |
| T88 | 5 | 5 | 0 | 0 |
| T89 | 5 | 1 | 2 | 2 |
| T91 | 4 | 5 | 0 | 0 |
| T92 | 5 | 0 | 3 | 2 |
| T94 | 5 | 0 | 5 | 0 |
| T96 | 5 | 0 | 5 | 0 |
| T97 | 5 | 0 | 2 | 3 |
| T98 | 5 | 0 | 1 | 4 |
| V01 | 5 | 0 | 3 | 2 |
| V02 | 4 | 0 | 2 | 3 |
| V03 | 5 | 0 | 5 | 0 |
| V04 | 0 | 0 | 5 | 0 |
| V05 | 5 | 0 | 5 | 0 |
| V06 | 5 | 0 | 5 | 0 |
| V07 | 5 | 0 | 5 | 0 |
| V08 | 5 | 5 | 0 | 0 |
| V09 | 5 | 5 | 0 | 0 |
| V10 | 5 | 0 | 0 | 5 |
| V11 | 5 | 0 | 5 | 0 |
| V12 | 5 | 0 | 1 | 4 |
| V13 | 5 | 0 | 1 | 4 |
| V14 | 5 | 0 | 5 | 0 |
| V15 | 5 | 5 | 0 | 0 |
| V16 | 5 | 0 | 0 | 5 |
| V17 | 5 | 0 | 5 | 0 |
| V18 | 5 | 0 | 3 | 2 |
| V19 | 5 | 0 | 5 | 0 |
| V20 | 3 | 0 | 0 | 5 |
| V21 | 5 | 0 | 0 | 5 |
| V22 | 5 | 0 | 4 | 1 |
| V23 | 5 | 0 | 2 | 3 |
| V24 | 5 | 0 | 5 | 0 |
| V25 | 5 | 0 | 0 | 5 |
| V26 | 5 | 0 | 0 | 5 |
| V27 | 5 | 0 | 0 | 5 |
| V28 | 5 | 0 | 1 | 4 |
| V29 | 5 | 0 | 2 | 3 |
| V30 | 5 | 0 | 4 | 1 |
| V31 | 5 | 0 | 3 | 2 |
| V32 | 0 | 1 | 1 | 2 |
| V33 | 5 | 0 | 4 | 1 |
| V34 | 5 | 0 | 2 | 3 |
| V35 | 5 | 0 | 3 | 2 |
| V36 | 5 | 0 | 0 | 5 |
| V37 | 5 | 0 | 2 | 3 |
| V38 | 5 | 0 | 0 | 5 |
| V39 | 5 | 0 | 0 | 5 |
| V40 | 5 | 0 | 0 | 5 |
| V41 | 5 | 0 | 0 | 5 |
| V42 | 2 | 0 | 0 | 5 |
| V43 | 5 | 0 | 5 | 0 |
| V44 | 5 | 0 | 5 | 0 |
| V45 | 5 | 1 | 4 | 0 |
| V46 | 5 | 0 | 1 | 4 |
| V47 | 5 | 5 | 0 | 0 |
| V48 | 5 | 0 | 0 | 5 |
| V49 | 5 | 0 | 1 | 4 |
| V52 | 5 | 0 | 0 | 5 |
| V54 | 5 | 0 | 1 | 4 |
| V56 | 0 | 0 | 5 | 0 |
| V58 | 0 | 0 | 1 | 4 |
| V60 | 3 | 0 | 0 | 5 |
| V62 | 0 | 0 | 0 | 5 |
| V66 | 2 | 0 | 0 | 3 |
| V67 | 5 | 0 | 0 | 5 |
| V68 | 5 | 0 | 0 | 5 |
| W01 | 4 | 0 | 0 | 5 |
| W02 | 5 | 0 | 0 | 5 |
| W03 | 5 | 0 | 0 | 5 |
| W04 | 5 | 0 | 0 | 5 |
| W05 | 5 | 0 | 5 | 0 |
| W06 | 5 | 0 | 0 | 5 |
| W07 | 3 | 0 | 0 | 5 |
| W08 | 5 | 0 | 2 | 3 |
| W09 | 5 | 0 | 5 | 0 |
| W10 | 5 | 0 | 3 | 2 |
| W11 | 3 | 0 | 3 | 2 |
| W12 | 5 | 0 | 0 | 5 |
| W13 | 5 | 0 | 1 | 4 |
| W14 | 5 | 0 | 1 | 4 |
| W15 | 5 | 0 | 0 | 5 |
| W16 | 5 | 0 | 0 | 5 |
| W17 | 5 | 0 | 0 | 5 |
| W18 | 5 | 0 | 4 | 1 |
| W19 | 5 | 5 | 0 | 0 |
| W20 | 5 | 5 | 0 | 0 |
| W21 | 5 | 5 | 0 | 0 |
| W22 | 5 | 5 | 0 | 0 |
| W23 | 5 | 5 | 0 | 0 |
| W24 | 5 | 5 | 0 | 0 |
| W25 | 5 | 5 | 0 | 0 |
| W26 | 5 | 5 | 0 | 0 |
| W27 | 5 | 0 | 0 | 5 |
| W28 | 5 | 0 | 5 | 0 |
| W29 | 5 | 0 | 5 | 0 |
| W30 | 5 | 0 | 5 | 0 |
| W31 | 5 | 0 | 5 | 0 |
| W32 | 5 | 0 | 4 | 1 |
| W33 | 5 | 0 | 5 | 0 |
| W34 | 5 | 0 | 3 | 2 |
| W37 | 5 | 0 | 1 | 4 |
| W38 | 5 | 0 | 0 | 5 |
| W39 | 5 | 0 | 0 | 5 |
| W40 | 5 | 0 | 0 | 5 |
| W41 | 5 | 0 | 0 | 5 |
| W42 | 5 | 0 | 0 | 5 |
| W43 | 5 | 0 | 0 | 5 |
| W44 | 5 | 0 | 0 | 5 |
| W45 | 5 | 0 | 0 | 5 |
| W46 | 5 | 0 | 0 | 5 |
| W47 | 5 | 0 | 0 | 5 |
| W48 | 5 | 0 | 2 | 3 |
| W49 | 5 | 0 | 5 | 0 |
| W50 | 5 | 0 | 0 | 5 |
| W51 | 5 | 0 | 1 | 4 |
| W52 | 5 | 0 | 0 | 5 |
| W53 | 5 | 0 | 0 | 5 |
| W54 | 5 | 0 | 0 | 5 |
| W55 | 5 | 0 | 0 | 5 |
| W56 | 5 | 0 | 1 | 4 |
| W57 | 5 | 0 | 2 | 3 |
| W58 | 5 | 0 | 0 | 5 |
| W59 | 5 | 0 | 1 | 4 |
| W60 | 5 | 0 | 0 | 5 |
| W61 | 5 | 0 | 0 | 5 |
| W62 | 5 | 0 | 0 | 5 |
| W63 | 5 | 0 | 0 | 5 |
| W64 | 5 | 0 | 1 | 4 |
| W65 | 5 | 5 | 0 | 0 |
| W67 | 5 | 5 | 0 | 0 |
| W68 | 5 | 5 | 0 | 0 |
| W69 | 5 | 0 | 4 | 1 |
| W70 | 5 | 0 | 4 | 1 |
| W71 | 5 | 0 | 2 | 3 |
| W72 | 5 | 0 | 0 | 5 |
| W73 | 5 | 0 | 1 | 4 |
| W74 | 5 | 0 | 0 | 5 |
| W75 | 5 | 0 | 5 | 0 |
| W76 | 5 | 0 | 5 | 0 |
| W77 | 5 | 0 | 0 | 5 |
| W78 | 5 | 0 | 3 | 2 |
| W79 | 5 | 0 | 0 | 5 |
| W80 | 5 | 0 | 2 | 3 |
| W81 | 5 | 0 | 4 | 1 |
| W82 | 5 | 0 | 5 | 0 |
| W83 | 5 | 0 | 3 | 2 |
| W84 | 5 | 0 | 1 | 4 |
| W85 | 5 | 0 | 5 | 0 |
| W86 | 5 | 0 | 3 | 2 |
| W87 | 5 | 0 | 4 | 1 |
| W88 | 5 | 0 | 1 | 4 |
| W89 | 5 | 0 | 4 | 1 |
| W91 | 5 | 0 | 4 | 1 |
| W92 | 5 | 0 | 5 | 0 |
| W93 | 5 | 0 | 0 | 5 |
| W94 | 5 | 0 | 0 | 5 |
| W95 | 5 | 0 | 0 | 5 |
| W96 | 5 | 0 | 0 | 5 |
| W97 | 5 | 0 | 0 | 5 |
| W98 | 5 | 0 | 0 | 5 |
| W99 | 5 | 5 | 0 | 0 |
| X01 | 5 | 4 | 1 | 0 |
| X02 | 5 | 2 | 1 | 2 |
| X03 | 5 | 1 | 2 | 1 |
| X05 | 2 | 1 | 3 | 1 |
| X07 | 5 | 0 | 5 | 0 |
| X08 | 5 | 0 | 5 | 0 |
| X09 | 5 | 0 | 5 | 0 |
| X10 | 5 | 0 | 5 | 0 |
| X11 | 5 | 0 | 5 | 0 |
| X12 | 5 | 0 | 5 | 0 |
| X14 | 5 | 3 | 2 | 0 |
| X15 | 5 | 0 | 0 | 5 |
| X16 | 4 | 0 | 1 | 4 |
| X19 | 2 | 0 | 0 | 5 |
| X20 | 4 | 0 | 1 | 4 |
| X21 | 5 | 0 | 0 | 5 |
| X22 | 5 | 0 | 0 | 5 |
| X23 | 3 | 0 | 0 | 5 |
| X24 | 1 | 0 | 0 | 5 |
| X25 | 5 | 0 | 0 | 5 |
| X27 | 5 | 0 | 0 | 5 |
| X45 | 5 | 4 | 1 | 0 |
| X46 | 5 | 2 | 3 | 0 |
| X53 | 5 | 3 | 2 | 0 |
| X55 | 5 | 5 | 0 | 0 |
| J77 | 1 | 0 | 1 | 0 |
| O20 | 1 | 0 | 0 | 2 |
| V57 | 0 | 0 | 1 | 1 |
| V61 | 0 | 0 | 0 | 2 |
| X04 | 0 | 0 | 0 | 1 |
| X17 | 0 | 2 | 0 | 0 |
| A82 | 0 | 0 | 2 | 0 |
| E66 | 2 | 0 | 2 | 0 |
| E67 | 1 | 1 | 1 | 0 |
| G12 | 2 | 1 | 1 | 0 |
| G20 | 2 | 1 | 1 | 0 |
| H37 | 0 | 0 | 2 | 0 |
| K73 | 2 | 0 | 2 | 0 |
| K74 | 2 | 0 | 2 | 0 |
| M57 | 2 | 0 | 1 | 1 |
| P28 | 2 | 0 | 1 | 1 |
| P30 | 2 | 0 | 1 | 1 |
| Q57 | 0 | 0 | 2 | 0 |
| T32 | 2 | 0 | 0 | 2 |
| V51 | 2 | 0 | 0 | 2 |

**Supplementary table 2. Contribution of each Office of Population Censuses and Surveys (OPCS) codes to each class of surgery, according to NHS England definitions.** Class1, emergency surgery; class 2, urgent surgery with a waiting time < 4 weeks; class 3, semi-urgent surgery with a waiting time < 3 months; class 4, elective surgery with a waiting time > 3 months. The count in each cell represents the number of years of data individual codes contributed data to that class. Where there is a 0, this means that there were no years of data where that code fitted the class definition.

| Category of surgery | Apr. | May | Jun. | Jul. | Aug. | Sept. | Oct. | Nov. | Dec. | Jan. | Feb. | Mar. |
| --- | --- | --- | --- | --- | --- | --- | --- | --- | --- | --- | --- | --- |
| Class 1 | 66 374 (1 144) | 69 379 (1 196) | 67 599 (1 165) | 69 044 (1 190) | 66 337 (1 143) | 67 157 (1 158) | 70 558 (1 216) | 69 833 (1 204) | 71 162 (1 227) | 71 091 (1 225) | 65 097 (1 122) | 71 896 (1 239) |
| Class 2 | 29 702 (1 821) | 30 891 (1 893) | 31 994 (1 961) | 32 069 (1 966) | 30 254 (1 854) | 31 274 (1 917) | 32 853 (2 014) | 32 684 (2 003) | 28 479 (1 746) | 31 389 (1 924) | 30 267 (1 855) | 32 692 (2 004) |
| Class 3 | 193 225 (32 211) | 200 961 (33 500) | 208 136 (34 696) | 208 628 (34 778) | 196 821 (32 810) | 203 455 (33 916) | 213 729 (35 629) | 212 630 (35 445) | 185 271 (30 885) | 204 205 (34 041) | 196 906 (32 824) | 212 682 (35 454) |
| Class 4 | 74 399 (23 502) | 77 378 (24 443) | 80 140 (25 316) | 80 330 (25 376) | 75 783 (23 940) | 78 338 (24 747) | 82 294 (25 996) | 81 871 (25 863) | 71 336 (22 535) | 78 627 (24 838) | 75 816 (23 950) | 81 891 (25 869) |

**Supplementary table 3.** The weighted five-year median number of procedures per month for each class of surgery. Numbers presented as n (Interquartile range). Procedures were divided by month according to NHS England monthly activity returns between 2014-2019.

| Category of surgery | Mar. | April | May | Jun. | Jul. | Aug. | Sep. | Oct. | Nov. | Dec. | Jan. | Feb. |
| --- | --- | --- | --- | --- | --- | --- | --- | --- | --- | --- | --- | --- |
| Class 1 | 72 999 (69 230 to 76 769) | 67 776 (63 324 to 72 227) | 70 844 (66 191 to 75 497) | 69 027 (64 493 to 73 561) | 70 502 (65 871 to 75 133) | 67 738 (63 289 to 72 187) | 68 575 (64 071 to 73 079) | 72 048 (67 316 to 76 781) | 71 308 (66 624 to 75 991) | 72 665 (67 892 to 77 438) | 72 593 (67 825 to 77 361) | 66 472 (62 106 to 70 838) |
| Class 2 | 35 290 (29 202 to 42 773) | 33 207 (26 706 to 41 786) | 34 536 (27 775 to 43 459) | 35 770 (28 767 to 45 010) | 35 854 (28 835 to 45 117) | 33 825 (27 203 to 42 563) | 34 965 (28 120 to 43 998) | 36 731 (29 540 to 46 220) | 36 542 (29 388 to 45 982) | 31 840 (25 607 to 40 066) | 35 094 (28 223 to 44 160) | 33 840 (27 215 to 42 582) |
| Class 3 | 168 255 (138 218 to 221 964) | 142 673 (117 536 to 200 739) | 148 386 (122 242 to 208 776) | 153 684 (126 607 to 216 230) | 154 047 (126 906 to 216 741) | 145 329 (119 724 to 204 475) | 150 228 (123 759 to 211 367) | 157 814 (130 009 to 222 041) | 157 002 (129 340 to 220 899) | 136 801 (112 698 to 192 476) | 150 781 (124 216 to 212 146) | 145 392 (119 776 to 204 563) |
| Class 4 | 116 037 (61 558 to 173 139) | 118 253 (55 674 to 184 595) | 122 988 (57 903 to 191 986) | 127 379 (59 970 to 198 840) | 127 680 (60 112 to 199 310) | 120 454 (56 710 to 188 030) | 124 514 (58 622 to 194 368) | 130 802 (61 582 to 204 183) | 130 129 (61 265 to 203 133) | 113 386 (53 382 to 176 996) | 124 973 (58 838 to 195 085) | 120 506 (56 735 to 188 111) |

**Supplementary table 4.** The estimated mean number (95% confidence interval) of surgical procedures per month for the period 1st March 2020 – 28th February 2021, assuming a pre-pandemic linear growth trajectory.

| Category of surgery | 0 to 14 years | 15 to 59 years | 60 to 74 years | 75+ years | Total procedures |
| --- | --- | --- | --- | --- | --- |
| Class1 | 5 707 (8.1%) | 41 394 (59.1%) | 11 516 (16.4%) | 11 431 (16.3%) | 70 048 |
| Class2 | 1 841 (5.4%) | 17 410 (50.8%) | 9 745 (28.4%) | 5 300 (15.5%) | 34 294 |
| Class3 | 7 274 (4.7%) | 64 668 (41.6%) | 43 010 (27.7%) | 40 332 (26.0%) | 155 284 |
| Class4 | 9 445 (8.0%) | 54 491 (46.4%) | 35 733 (30.4%) | 17 856 (15.2%) | 117 527 |

**Supplementary table 5.** Proportion of patients in each age class from an average forecasted month from 2019-2020 and 2020-2021 across classes.

| Class | Mar | April | May | Jun. | Jul. | Aug. | Sep. | Oct. | Nov. | Dec. | Jan. | Feb. |
| --- | --- | --- | --- | --- | --- | --- | --- | --- | --- | --- | --- | --- |
| Class 1 | 0 (0 to 0) | 0 (0 to 0) | 0 (0 to 0) | 0 (0 to 0) | 0 (0 to 0) | 0 (0 to 0) | 0 (0 to 0) | 0 (0 to 0) | 0 (0 to 0) | 0 (0 to 0) | 0 (0 to 0) | 0 (0 to 0) |
| Class 2 | 7 058 (5 841 to 8 555) | 40 265 (32 546 to 50 341) | 74 801 (60 321 to 938 00) | 110 571 (89 088 to 138 810) | 146 425 (117 923 to 183 927) | 180 250 (145 126 to 226 490) | 215 215 (173 246 to 270 488) | 251 946 (202 786 to 316 708) | 288 488 (232 174 to 362 690) | 320 328 (257 781 to 402 756) | 355 422 (286 004 to 446 916) | 389 262 (313 219 to 489 498) |
| Class 3 | 84 128 (69 109 to 110 982) | 226 801 (186 645 to 311 721) | 375 187 (308 887 to 520 497) | 528 871 (435 494 to 736 727) | 682 918 (562 400 to 953 468) | 828 247 (682 124 to 1 157 943) | 978 475 (805 883 to 1 369 310) | 1 136 289 (935 892 to 1 591 351) | 1 293 291 (1 065 232 to 1 812 250) | 1 430 092 (1 177 930 to 2 004 726) | 1 580 873 (1 302 146 to 2 216 872) | 1 726 265 (1 421 922 to 2 421 435) |
| Class 4 | 58 018 (30 779 to 86 570) | 176 271 (86 453 to 271 165) | 299 259 (144 356 to 463 151) | 426 638 (204 326 to 661 991) | 554 318 (264 438 to 861 301) | 674 772 (321 148 to 1 049 331) | 799 286 (379 770 to 1 243 699) | 930 088 (441 352 to 1 447 882) | 1 060 217 (502 617 to 1 651 015) | 1 173 603 (555 999 to 1 828 011) | 1 298 576 (614 837 to 2 023 096) | 1 419 082 (671 572 to 2 211 207) |
| Total | 149 204 (105 728 to 206 107) | 443 337 (305 644 to 633 227) | 749 247 (513 564 to 1 077 448) | 1 066 080 (728 908 to 1 537 528) | 1 383 661 (944 761 to 1 998 696) | 1 683 269 (1 148 398 to 2 433 764) | 1 992 976 (1 358 899 to 2 883 497) | 2 318 323 (1 580 030 to 3 355 941) | 2 641 996 (1 800 023 to 3 825 955) | 2 924 023 (1 991 710 to 4 235 493) | 3 234 871 (2 202 987 to 4 686 884) | 3 534 609 (2 406 713 to 5 122 140) |

**Supplementary table 6** Cumulative deficit of surgical activity on the last day of each month, from 1^st^ March 2020 to 28^th^ February 2021, if only class 1 (emergency) surgical activity continued at pre-pandemic levels, class 2 activity occurred at 80% capacity in March and class 3 and 4 activity occurred at 50% capacity in March. Numbers are presented as time-weighted average with 95% confidence intervals).

| Scenario | Mar | April | May | Jun. | Jul. | Aug. | Sep. | Oct. | Nov. | Dec. | Jan. | Feb. |
| --- | --- | --- | --- | --- | --- | --- | --- | --- | --- | --- | --- | --- |
| 20% | 7 058 (5 840 to 8 555) | 33 624 (27 205 to 41 984) | 61 253 (49 425 to 76 751) | 82 715 (66 686 to 103 757) | 97 056 (78 220 to 121 805) | 103 821 (83 659 to 130 316) | 103 821 (83 659 to 130 314) | 103 822 (83 659 to 130 314) | 103 824 (83 657 to 130 316) | 103 824 (83 659 to 130 317) | 103 823 (83 657 to 130 317) | 103 823 (83 657 to 130 319) |
| 40% | 7 058 (5 840 to 8 555) | 26 982 (21 864 to 33 627) | 47 704 (38 529 to 59 702) | 63 800 (51 474 to 79 956) | 74 556 (60 125 to 93 491) | 79 630 (64 205 to 99 875) | 79 630 (64 205 to 99 876) | 79 631 (64 205 to 99 876) | 79 631 (64 205 to 99 876) | 79 631 (64 204 to 99 877) | 79 630 (64 205 to 99 877) | 79 630 (64 205 to 99 877) |
| 60% | 7 058 (5 840 to 8 555) | 20 341 (16 522 to 25 269) | 34 155 (27 632 to 42 653) | 44 886 (36 262 to 56 156) | 52 058 (42 029 to 65 179) | 55 440 (44 749 to 69 435) | 55 440 (44 749 to 69 435) | 55 439 (44 749 to 69 435) | 55 439 (44 749 to 69 435) | 55 439 (44 749 to 69 434) | 55 440 (44 749 to 69 434) | 55 440 (44 749 to 69 434) |
| 80% | 7 058 (5 840 to 8 555) | 13 699 (11 181 to 16 912) | 20 606 (16 736 to 25 604) | 25 972 (21 051 to 32 356) | 29 557 (23 934 to 36 867) | 31 248 (25 295 to 38 996) | 31 248 (25 295 to 38 996) | 31 248 (25 295 to 38 996) | 31 248 (25 295 to 38 996) | 31 248 (25 294 to 38 996) | 31 248 (25 295 to 38 996) | 31 248 (25 295 to 38 996) |

**Supplementary table 7.** Cumulative monthly deficit of class 2 surgical activity between 1^st^ March 2020 and 28^th^ February 2021, for four scenarios of class 2 activity (20% - 80% of time-weighted pre-pandemic activity) and assuming activity increases linearly from 1^st^ June to reach pre-pandemic expected levels by 31^st^ August.

| Item | Mar | April | May | Jun. | Jul. | Aug. | Sep. | Oct. | Nov. | Dec. | Jan. | Feb. |
| --- | --- | --- | --- | --- | --- | --- | --- | --- | --- | --- | --- | --- |
| Deficit of admissions | 149 204 (105 728 to 206 106) | 416 771 (284 279 to 599 797) | 695 052 (469 979 to 1 009 251) | 981 481 (660 871 to 1 431 073) | 1 266 793 (850 772 to 1 851 635) | 1 534 267 (1 028 567 to 2 246 269) | 1 771 452 (1 180 008 to 2 599 162) | 1 981 161 (1 306 595 to 2 914 366) | 2 150 540 (1 400 195 to 3 172 724) | 2 235 580 (1 440 230 to 3 305 471) | 2 298 067 (1 469 650 to 3 403 014) | 2 328 193 (1 483 834 to 3 450 043) |
| Bed days | 210 547 (143 876 to 292 608) | 584 294 (379 079 to 846 717) | 973 006 (623 700 to 1 423 014) | 1 369 659 (872 278 to 2 012 410) | 1 761 292 (1 116 653 to 2 595 703) | 2 125 143 (1 342 685 to 3 138 922) | 2 456 761 (1 539 788 to 3 638 701) | 2 764 480 (1 713 360 to 4 106 531) | 3 030 177 (1 852 726 to 4 515 062) | 3 177 387 (1 922 027 to 4 744 855) | 3 285 556 (1 972 957 to 4 913 708) | 3 337 706 (1 997 510 to 4 995 117) |
| Day cases | 96 974 (71 054 to 132 677) | 268 910 (191 293 to 383 405) | 447 731 (316 346 to 644 171) | 632 039 (445 142 to 913 117) | 815 881 (573 518 to 1 181 565) | 988 471 (693 947 to 1 433 754) | 1 137 530 (794 275 to 1 653 281) | 1 264 209 (875 032 to 1 841 815) | 1 360 482 (930 861 to 1 987 516) | 1 403 953 (951 326 to 2 055 374) | 1 435 895 (966 365 to 2 105 236) | 1 451 295 (973 616 to 2 129 276) |
| PPE Items (low) | 2 387 264 (1 691 648 to 3 297 696) | 6 668 336 (4 548 464 to 9 596 752) | 11 120 832 (7 519 664 to 16 148 016) | 15 703 696 (10 573 936 to 22 897 168) | 20 268 688 (13 612 352 to 29 626 160) | 24 548 272 (16 457 072 to 35 940 304) | 28 343 232 (18 880 128 to 41 586 592) | 31 698 576 (20 905 520 to 46 629 856) | 34 408 640 (22 403 120 to 50 763 584) | 35 769 280 (23 043 680 to 52 887 536) | 36 769 072 (23 514 400 to 54 448 224) | 37 251 088 (23 741 344 to 55 200 672) |
| PPE Items  (high) | 4 774 528 (3 383 296 to 6 595 392) | 13 336 672 (9 096 928 to 19 193 504) | 22 241 664 (15 039 328 to 32 296 032) | 31 407 392 (21 147 872 to 45 794 336) | 40 537 376 (27 224 704 to 59 252 320) | 49 096 544 (32 914 144 to 71 880 608) | 56 686 464 (37 760 256 to 83 173 184) | 63 397 152 (41 811 040 to 93 259 712) | 68 817 280 (44 806 240 to 101 527 168) | 71 538 560 (46 087 360 to 105 775 072) | 73 538 144 (47 028 800 to 108 896 448) | 74 502 176 (47 482 688 to 110 401 344) |
| Crit care admissions | 523 (346 to 735) | 1 479 (931 to 2 163) | 2 474 (1 536 to 3 650) | 3 494 (2 158 to 5 180) | 4 509 (2 773 to 6 701) | 5 458 (3 346 to 8 125) | 6 340 (3 857 to 9 458) | 7 169 (4 315 to 10 725) | 7 901 (4 694 to 11 851) | 8 316 (4 890 to 12 500) | 8 622 (5 033 to 12 977) | 8 769 (5 103 to 13 207) |
| SARS-CoV 2 PCR | 298 408 (211 456 to 412 212) | 833 542 (568 558 to 1 199 594) | 1 390 104 (939 958 to 2 018 502) | 1 962 962 (1 321 742 to 2 862 146) | 2 533 586 (1 701 544 to 3 703 270) | 3 068 534 (2 057 134 to 4 492 538) | 3 542 904 (2 360 016 to 5 198 324) | 3 962 322 (2 613 190 to 5 828 732) | 4 301 080 (2 800 390 to 6 345 448) | 4 471 160 (2 880 460 to 6 610 942) | 4 596 134 (2 939 300 to 6 806 028) | 4 656 386 (2 967 668 to 6 900 084) |
| Excess bed days for screening | 52 230 (34 674 to 73 429) | 147 861 (92 986 to 216 392) | 247 321 (153 633 to 365 080) | 349 442 (215 729 to 517 956) | 450 912 (277 254 to 670 070) | 545 796 (334 620 to 812 515) | 633 922 (385 733 to 945 881) | 716 952 (431 563 to 1 072 551) | 790 058 (469 334 to 1 185 208) | 831 627 (488 904 to 1 250 097) | 862 172 (503 285 to 1 297 778) | 876 898 (510 218 to 1 320 766) |
| Excess cost (low estimate) [£] | 24 910 972 (17 082 910 to 34 723 156) | 70 063 136 (45 872 377 to 101 708 323) | 117 023 296 (75 814 543 to 171 375 534) | 165 297 216 (106 533 783 to 243 073 182) | 213 321 076 (137 032 888 to 314 482 134) | 258 283 791 (165 530 099 to 381 418 042) | 299 126 133 (190 351 633 to 442 732 953) | 336 483 165 (211 856 355 to 499 338 615) | 368 128 662 (228 703 500 to 547 887 644) | 385 202 347 (236 741 421 to 574 539 578) | 397 748 011 (242 648 148 to 594 123 520) | 403 796 481 (245 495 903 to 603 565 443) |
| Excess cost (high estimate) [£] | 37 980 082 (26 025 847 to 52 950 638) | 106 836 811 (69 884 558 to 155 120 983) | 178 450 717 (115 499 331 to 261 382 165) | 252 066 043 (162 295 806 to 370 737 845) | 325 297 967 (208 754 801 to 479 650 714) | 393 859 952 (252 162 301 to 581 738 944) | 456 171 658 (289 990 315 to 675 302 953) | 513 206 941 (322 789 665 to 761 740 798) | 561 568 897 (348 516 755 to 835 943 901) | 587 697 548 (360 817 557 to 876 730 589) | 606 896 760 (369 856 894 to 906 700 804) | 616 153 013 (374 214 946 to 921 150 217) |

**Supplementary table 8.** Resource requirements associated with the cumulative monthly deficit of surgical activity between 1^st^ March 2020 and 28^th^ February 2021, reported on the last day of each month. Total number of procedures is presented as a time-weighted average with 95% confidence intervals derived from a linear growth model at procedure group level. Costs are presented in pounds sterling and include only excess costs associated with specific measures required for surgical time. Low estimate: four members of staff in personal protective equipment (PPE), bed day cost of £222 (€243.68). High estimate: eight members of staff in personal protective equipment, bed day cost of £346 (€379.79). Full cost data is in supplementary table 10. Costs are provided in Euros (€).

| Item | Mar | April | May | Jun. | Jul. | Aug. | Sep. | Oct. | Nov. | Dec. | Jan. | Feb. | Total |
| --- | --- | --- | --- | --- | --- | --- | --- | --- | --- | --- | --- | --- | --- |
| Total admissions | 243 377 (192 480 to 308 539) | 94 342 (84 689 to 105 656) | 98 473 (88 411 to 110 264) | 99 431 (88 945 to 111 819) | 102 771 (91 823 to 115 739) | 99 872 (89 131 to 112 621) | 141 097 (123 131 to 169 919) | 187 686 (161 860 to 234 021) | 225 602 (193 017 to 287 647) | 269 652 (219 544 to 354 229) | 320 954 (249 682 to 431 209) | 336 084 (251 648 to 459 066) | 2 219 341 (1 834 361 to 2 800 729) |
| Cost of PCR tests | 10 151 517 (8 028 548 to 12 869 495) | 3 935 107 (3 532 469 to 4 407 026) | 4 107 415 (3 687 718 to 4 599 230) | 4 147 375 (3 709 992 to 4 664 091) | 4 286 690 (3 830 037 to 4 827 599) | 4 165 769 (3 717 751 to 4 697 544) | 5 885 308 (5 135 928 to 7 087 505) | 7 828 586 (6 751 355 to 9 761 268) | 9 410 103 (8 050 947 to 11 998 067) | 11 247 476 (9 157 417 to 14 775 275) | 13 387 338 (10 414 506 to 17 986 193) | 14 018 426 (10 496 509 to 19 148 138) | 92 571 110 (76 513 177 to 116 821 431) |
| Cost of CT scans | 1 788 111 (1 695 786 to 1 880 512) | 1 660 189 (1 551 125 to 1 769 252) | 1 735 321 (1 621 335 to 1 849 308) | 1 690 863 (1 579 754 to 1 801 896) | 1 726 990 (1 613 534 to 1 840 371) | 1 659 280 (1 550 292 to 1 768 268) | 1 679 729 (1 569 454 to 1 790 080) | 1 764 859 (1 648 904 to 1 880 739) | 1 746 682 (1 631 938 to 1 861 426) | 1 779 931 (1 663 067 to 1 896 872) | 1 778 189 (1 661 401 to 1 894 978) | 1 628 227 (1 521 284 to 1 735 170) | 20 638 371 (19 307 874 to 21 968 872) |
| Cost of excess bed days for isolation (low estimate) | 33 084 993 (27 445 556 to 39 714 266) | 19 739 162 (18 023 216 to 21 656 528) | 20 615 757 (18 825 592 to 22 615 714) | 20 509 702 (18 682 650 to 22 567 541) | 21 095 427 (19 200 449 to 23 235 509) | 20 405 513 (18 557 918 to 22 496 756) | 23 167 309 (20 849 080 to 26 262 083) | 26 665 851 (23 820 504 to 30 863 923) | 28 752 860 (25 520 183 to 33 864 884) | 33 012 555 (27 885 469 to 40 350 606) | 38 280 687 (30 779 949 to 48 336 338) | 39 641 833 (30 566 597 to 51 294 650) | 324 971 649 (280157163 to 383258798) |
| Cost of gloves (low estimate) | 318 437 (251 843 to 403 695) | 123 438 (110 808 to 138 242) | 128 843 (115 678 to 144 271) | 130 097 (116 376 to 146 305) | 134 467 (120 142 to 151 434) | 130 673 (116 620 to 147 354) | 184 613 (161 106 to 222 323) | 245 570 (211 779 to 306 196) | 295 180 (252 546 to 376 360) | 352 816 (287 254 to 463 478) | 419 938 (326 687 to 564 198) | 439 736 (329 258 to 600 647) | 2 903 808 (2 400 097 to 3 664 503) |
| Cost of FFP3 (low estimate) | 3 098 885 (2 450 821 to 3 928 583) | 1 201 243 (1 078 332 to 1 345 303) | 1 253 842 (1 125 724 to 1 403 975) | 1 266 041 (1 132 524 to 1 423 776) | 1 308 569 (1 169 169 to 1 473 688) | 1 271 656 (1 134 892 to 1 433 987) | 1 796 568 (1 567 810 to 2 163 554) | 2 389 779 (2 060 941 to 2 979 756) | 2 872 557 (2 457 658 to 3 662 567) | 3 433 440 (2 795 422 to 4 510 347) | 4 086 661 (3 179 165 to 5 490 521) | 4 279 309 (3 204 199 to 5 845 221) | 28 258 550 (23 356 657 to 35 661 278) |
| Cost of gowns (low estimate) | 3 205 742 (2 535 331 to 4 064 051) | 1 242 665 (1 115 517 to 1 391 692) | 1 297 079 (1 164 543 to 1 452 388) | 1 309 697 (1 171 577 to 1 472 871) | 1 353 692 (1 209 486 to 1 524 505) | 1 315 506 (1 174 027 to 1 483 434) | 1 858 518 (1 621 871 to 2 238 160) | 2 472 185 (2 132 007 to 3 082 506) | 2 971 611 (2 542 404 to 3 788 863) | 3 551 835 (2 891 816 to 4 665 875) | 4 227 580 (3 288 791 to 5 679 850) | 4 426 872 (3 314 687 to 6 046 780) | 29 232 982 (24 162 057 to 36 890 975) |
| Cost of eye protection (low estimate | 3 098 885 (2 450 821 to 3 928 583) | 1 201 243 (1 078 332 to 1 345 303) | 1 253 842 (1 125 724 to 1 403 975) | 1 266 041 (1 132 524 to 1 423 776) | 1 308 569 (1 169 169 to 1 473 688) | 1 271 656 (1 134 892 to 1 433 987) | 1 796 568 (1 567 810 to 2 163 554) | 2 389 779 (2 060 941 to 2 979 756) | 2 872 557 (2 457 658 to 3 662 567) | 3 433 440 (2 795 422 to 4 510 347) | 4 086 661 (3 179 165 to 5 490 521) | 4 279 309 (3 204 199 to 5 845 221) | 28 258 550 (23 356 657 to 35 661 278) |
| Excess cost  (low estimate) | 54 746 570 (44 858 704 to 66 789 186) | 29 103 048 (26 489 800 to 32 053 345) | 30 392 099 (27 666 315 to 33 468 862) | 30 319 814 (27 525 397 to 33 500 255) | 31 214 402 (28 311 986 to 34 526 794) | 30 220 053 (27 386 392 to 33 461 331) | 36 368 613 (32 473 059 to 41 927 260) | 43 756 608 (38 686 430 to 51 854 143) | 48 921 551 (42 913 333 to 59 214 737) | 56 811 493 (47 475 867 to 71 172 800) | 66 267 055 (52 829 664 to 85 442 602) | 68 713 711 (52 636 732 to 90 515 828) | 526 835 017 (449 253 679 to 633 927 143) |
| Cost of excess bed days for isolation (high estimate) | 51 564 899 (42 775 508 to 61 897 008) | 30 764 640 (28 090 238 to 33 752 967) | 32 130 863 (29 340 789 to 35 247 915) | 31 965 572 (29 118 004 to 35 172 833) | 32 878 458 (29 925 024 to 36 213 900) | 31 803 187 (28 923 603 to 35 062 511) | 36 107 606 (32 494 510 to 40 930 994) | 41 560 291 (37 125 651 to 48 103 230) | 44 813 016 (39 774 700 to 52 780 406) | 51 452 000 (43 461 136 to 62 888 782) | 59 662 694 (47 972 355 to 75 335 013) | 61 784 117 (47 639 832 to 79 945 715) | 506 487 343 (436 641 350 to 597 331 274) |
| Cost of gloves (high estimate) | 636 874 (503 686 to 807 391) | 246 876 (221 616 to 276 483) | 257 686 (231 356 to 288 541) | 260 193 (232 754 to 292 610) | 268 933 (240 285 to 302 869) | 261 347 (233 240 to 294 709) | 369 226 (322 211 to 444 648) | 491 141 (423 559 to 612 391) | 590 360 (505 091 to 752 721) | 705 631 (574 508 to 926 954) | 839 880 (653 373 to 1 128 397) | 879 471 (658 517 to 1 201 293) | 5 807 618 (4 800 196 to 7 329 007) |
| Cost of FFP3 (high estimate) | 6 197 769 (4 901 640 to 7 857 166) | 2 402 486 (2 156 666 to 2 690 605) | 2 507 685 (2 251 449 to 2 807 951) | 2 532 082 (2 265 048 to 2 847 550) | 2 617 137 (2 338 338 to 2 947 376) | 2 543 311 (2 269 784 to 2 867 974) | 3 593 136 (3 135 619 to 4 327 108) | 4 779 558 (4 121 880 to 5 959 511) | 5 745 116 (4 915 315 to 7 325 135) | 6 866 880 (5 590 844 to 9 020 693) | 8 173 322 (6 358 330 to 10 981 044) | 8 558 619 (6 408 396 to 11 690 442) | 56 517 101 (46 713 309 to 71 322 555) |
| Cost of gowns (high estimate) | 6 411 485 (5 070 662 to 8 128 102) | 2 485 330 (2 231 033 to 2 783 385) | 2 594 157 (2 329 085 to 2 904 777) | 2 619 394 (2 343 153 to 2 945 742) | 2 707 382 (2 418 970 to 3 049 010) | 2 631 012 (2 348 053 to 2 966 870) | 3 717 037 (3 243 743 to 4 476 318) | 4 944 370 (4 264 014 to 6 165 011) | 5 943 224 (5 084 809 to 7 577 726) | 7 103 669 (5 783 632 to 9 331 752) | 8 455 161 (6 577 582 to 11 359 701) | 8 853 744 (6 629 376 to 12 093 561) | 58 465 965 (48 324 112 to 73 781 955) |
| Cost of eye protection (high estimate | 6 197 769 (4 901 640 to 7 857 166) | 2 402 486 (2 156 666 to 2 690 605) | 2 507 685 (2 251 449 to 2 807 951) | 2 532 082 (2 265 048 to 2 847 550) | 2 617 137 (2 338 338 to 2 947 376) | 2 543 311 (2 269 784 to 2 867 974) | 3 593 136 (3 135 619 to 4 327 108) | 4 779 558 (4 121 880 to 5 959 511) | 5 745 116 (4 915 315 to 7 325 135) | 6 866 880 (5 590 844 to 9 020 693) | 8 173 322 (6 358 330 to 10 981 044) | 8 558 619 (6 408 396 to 11 690 442) | 56 517 101 (46 713 309 to 71 322 555) |
| Excess cost (high estimate) | 82 948 422 (67 877 470 to 101 296 839) | 43 897 115 (39 939 812 to 48 370 324) | 45 840 812 (41 713 181 to 50 505 674) | 45 747 560 (41 513 751 to 50 572 273) | 47 102 728 (42 704 526 to 52 128 500) | 45 607 218 (41 312 507 to 50 525 850) | 54 945 177 (49 037 084 to 63 383 763) | 66 148 362 (58 457 243 to 78 441 662) | 73 993 616 (64 878 116 to 89 620 617) | 86 022 467 (71 821 448 to 107 861 022) | 100 469 906 (79 995 877 to 129 666 370) | 104 281 222 (79 762 308 to 137 504 764) | 797 004 605 (679 013 323 to 959 877 658) |
| Cost of surgical procedure | 546 368 931 (421 484 207 to 695 212 910) | 233 617 851 (209 490 535 to 261 965 892) | 243 838 357 (218 689 876 to 273 380 264) | 246 436 382 (220 199 785 to 277 506 001) | 254 790 483 (227 388 674 to 287 325 154) | 247 673 121 (220 780 904 to 279 667 708) | 311 483 911 (273 438 015 to 367 683 110) | 384 559 887 (334 490 319 to 466 928 084) | 438 703 011 (378 901 606 to 543 792 703) | 542 518 796 (435 112 327 to 703 562 535) | 673 009 515 (506 387 039 to 901 714 778) | 730 401 779 (522 628 771 to 1 000 899 463) | 4 853 402 024 (3 968 992 057 to 6 059 638 601) |
| Grand total (low estimate) | 601 115 501 (466 342 911 to 762 002 096) | 262 720 899 (235 980 335 to 294 019 237) | 274 230 456 (246 356 191 to 306 849 126) | 276 756 196 (247 725 182 to 311 006 256) | 286 004 885 (255 700 660 to 321 851 948) | 277 893 174 (248 167 296 to 313 129 039) | 347 852 524 (305 911 074 to 409 610 370) | 428 316 495 (373 176 749 to 518 782 227) | 487 624 562 (421 814 939 to 603 007 440) | 599 330 289 (482 588 194 to 774 735 335) | 739 276 570 (559 216 703 to 987 157 380) | 799 115 490 (575 265 503 to 1 091 415 291) | 5 380 237 041 (4 418 245 737 to 6 693 565 745) |
| Grand total (high estimate) | 629 317 353 (489 361 677 to 796 509 749) | 277 514 966 (249 430 347 to 310 336 216) | 289 679 169 (260 403 057 to 323 885 938) | 292 183 942 (261 713 536 to 328 078 274) | 301 893 211 (270 093 200 to 339 453 654) | 293 280 339 (262 093 411 to 330 193 558) | 366 429 088 (322 475 099 to 431 066 873) | 450 708 249 (392 947 562 to 545 369 746) | 512 696 627 (443 779 722 to 633 413 320) | 628 541 263 (506 933 775 to 811 423 557) | 773 479 421 (586 382 916 to 1 031 381 148) | 834 683 001 (602 391 079 to 1 138 404 227) | 5 650 406 629 (4 648 005 381 to 7 019 516 260) |

**Supplementary table 9.** Cost of resources for reintroduction of surgical activity between 1^st^ March 2020 and 28^th^ February 2021, reported on the last day of each month. Total number of procedures is presented as a time-weighted average with 95% confidence intervals derived from a linear growth model at procedure group level. Costs are presented in pounds sterling and include only excess costs associated with specific measures required for surgical time. Low estimate: four members of staff in personal protective equipment, bed day cost of £222 (€243.68). High estimate: eight members of staff in personal protective equipment, bed day cost of £346 (€379.79). Costs are provided in Euros (€). These estimates include the continuation of class 1 surgical activity.

| Item | Mar | April | May | Jun. | Jul. | Aug. | Sep. | Oct. | Nov. | Dec. | Jan. | Feb. |
| --- | --- | --- | --- | --- | --- | --- | --- | --- | --- | --- | --- | --- |
| Deficit of admissions | 149 204 (105 728 to 206 106) | 416 771 (284 279 to 599 797) | 695 052 (469 979 to 1 009 251) | 981 481 (660 871 to 1 431 073) | 1 266 793 (850 772 to 1 851 635) | 1 534 267 (1 028 567 to 2 246 269) | 1 771 452 (1 180 008 to 2 599 162) | 1 981 161 (1 306 595 to 2 914 366) | 2 150 540 (1 400 195 to 3 172 724) | 2 235 580 (1 440 230 to 3 305 471) | 2 298 067 (1 469 650 to 3 403 014) | 2 328 193 (1 483 834 to 3 450 042) |
| Cost of PCR tests | 6 223 460 (4 410 029 to 8 596 903) | 17 383 968 (11 857 585 to 25 018 181) | 28 991 370 (19 603 332 to 42 096 949) | 40 938 632 (27 565 643 to 59 691 601) | 52 839 304 (35 486 619 to 77 233 695) | 63 995 934 (42 902 641 to 93 694 306) | 73 889 176 (49 219 408 to 108 413 854) | 82 636 365 (54 499 489 to 121 561 353) | 89 701 346 (58 403 646 to 132 337 744) | 93 248 456 (60 073 549 to 137 874 765) | 95 854 856 (61 300 689 to 141 943 389) | 97 111 444 (61 892 319 to 143 904 977) |
| Cost of excess bed days for isolation (low estimate) | 12 727 403 (8 449 466 to 17 893 134) | 36 030 816 (22 658 971 to 52 730 626) | 60 267 349 (37 437 410 to 88 963 002) | 85 152 301 (52 568 959 to 126 215 878) | 109 878 405 (67 561 294 to 163 282 948) | 132 999 962 (81 540 277 to 197 994 178) | 154 474 459 (93 995 575 to 230 492 863) | 174 707 258 (105 163 566 to 261 359 873) | 192 521 753 (114 367 605 to 288 812 131) | 202 651 317 (119 136 383 to 304 624 327) | 210 094 473 (122 640 757 to 316 243 188) | 213 682 940 (124 330 288 to 321 844 941) |
| Cost of gloves (low estimate) | 195 221 (138 336 to 269 671) | 545 308 (371 953 to 784 781) | 909 413 (614 926 to 1 320 515) | 1 284 180 (864 690 to 1 872 432) | 1 657 486 (1 113 160 to 2 422 699) | 2 007 451 (1 345 787 to 2 939 043) | 2 317 787 (1 543 935 to 3 400 772) | 2 592 172 (1 709 563 to 3 813 188) | 2 813 790 (1 832 029 to 4 151 226) | 2 925 057 (1 884 412 to 4 324 914) | 3 006 816 (1 922 905 to 4 452 540) | 3 046 233 (1 941 464 to 4 514 072) |
| Cost of FFP3 (low estimate) | 1 899 793 (1 346 220 to 2 624 319) | 5 306 685 (3 619 684 to 7 637 129) | 8 849 996 (5 984 175 to 12 850 647) | 12 497 057 (8 414 775 to 18 221 646) | 16 129 893 (10 832 757 to 23 576 602) | 19 535 600 (13 096 596 to 28 601 419) | 22 555 643 (15 024 873 to 33 094 755) | 25 225 837 (16 636 686 to 37 108 202) | 27 382 516 (17 828 482 to 40 397 838) | 28 465 318 (18 338 242 to 42 088 086) | 29 260 956 (18 712 842 to 43 330 087) | 29 644 546 (18 893 445 to 43 928 888) |
| Cost of gowns (low estimate) | 1 965 303 (1 392 641 to 2 714 812) | 5 489 674 (3 744 500 to 7 900 479) | 9 155 170 (6 190 526 to 13 293 774) | 12 927 989 (8 704 940 to 18 849 979) | 16 686 096 (11 206 301 to 24 389 588) | 20 209 242 (13 548 203 to 29 587 676) | 23 333 423 (15 542 971 to 34 235 954) | 26 095 694 (17 210 365 to 38 387 796) | 28 326 741 (18 443 257 to 41 790 866) | 29 446 881 (18 970 595 to 43 539 399) | 30 269 955 (19 358 113 to 44 824 228) | 30 666 772 (19 544 943 to 45 443 677) |
| Cost of eye protection (low estimate) | 1 899 793 (1 346 220 to 2 624 319) | 5 306 685 (3 619 684 to 7 637 129) | 8 849 996 (5 984 175 to 12 850 647) | 12 497 057 (8 414 775 to 18 221 646) | 16 129 893 (10 832 757 to 23 576 602) | 19 535 600 (13 096 596 to 28 601 419) | 22 555 643 (15 024 873 to 33 094 755) | 25 225 837 (16 636 686 to 37 108 202) | 27 382 516 (17 828 482 to 40 397 838) | 28 465 318 (18 338 242 to 42 088 086) | 29 260 956 (18 712 842 to 43 330 087) | 29 644 546 (18 893 445 to 43 928 888) |
| Excess cost (low estimate) | 24 910 972 (17 082 910 to 34 723 156) | 70 063 136 (45 872 377 to 101 708 323) | 117 023 296 (75 814 543 to 171 375 534) | 165 297 216 (106 533 783 to 243 073 182) | 213 321 076 (137 032 888 to 314 482 134) | 258 283 791 (165 530 099 to 381 418 042) | 299 126 133 (190 351 633 to 442 732 953) | 336 483 165 (211 856 355 to 499 338 615) | 368 128 662 (228 703 500 to 547 887 644) | 385 202 347 (236 741 421 to 574 539 578) | 397 748 011 (242 648 148 to 594 123 520) | 403 796 481 (245 495 903 to 603 565 443) |
| Cost of excess bed days for isolation (high estimate) | 19 836 404 (13 168 988 to 27 887 497) | 56 156 138 (35 315 332 to 82 183 769) | 93 930 194 (58 348 396 to 138 654 049) | 132 714 848 (81 931 801 to 196 714 838) | 171 251 929 (105 298 233 to 254 486 037) | 207 288 228 (127 085 298 to 308 585 523) | 240 757 488 (146 497 608 to 359 236 626) | 272 291 492 (163 903 577 to 407 344 667) | 300 056 426 (178 248 611 to 450 130 619) | 315 843 944 (185 681 030 to 474 774 852) | 327 444 539 (191 142 802 to 492 883 529) | 333 037 375 (193 776 035 to 501 614 189) |
| Cost of gloves (high estimate) | 390 440 (276 671 to 539 343) | 1 090 615 (743 908 to 1 569 562) | 1 818 827 (1 229 851 to 2 641 030) | 2 568 360 (1 729 382 to 3 744 863) | 3 314 971 (2 226 318 to 4 845 398) | 4 014 902 (2 691 576 to 5 878 085) | 4 635 573 (3 087 869 to 6 801 544) | 5 184 344 (3 419 125 to 7 626 376) | 5 627 579 (3 664 060 to 8 302 453) | 5 850 114 (3 768 825 to 8 649 828) | 6 013 631 (3 845 811 to 8 905 081) | 6 092 465 (3 882 928 to 9 028 145) |
| Cost of FFP3 (high estimate) | 3 799 586 (2 692 439 to 5 248 636) | 10 613 370 (7 239 367 to 15 274 257) | 17 699 994 (11 968 349 to 25 701 296) | 24 994 112 (16 829 551 to 36 443 294) | 32 259 786 (21 665 514 to 47 153 203) | 39 071 202 (26 193 190 to 57 202 840) | 45 111 287 (30 049 743 to 66 189 511) | 50 451 676 (33 273 372 to 74 216 406) | 54 765 032 (35 656 963 to 80 795 676) | 56 930 636 (36 676 483 to 84 176 173) | 58 521 912 (37 425 684 to 86 660 175) | 59 289 092 (37 786 889 to 87 857 776) |
| Cost of gowns (high estimate) | 3 930 606 (2 785 281 to 5 429 624) | 10 979 348 (7 489 000 to 15 800 956) | 18 310 339 (12 381 052 to 26 587 547) | 25 855 979 (17 409 879 to 37 699 958) | 33 372 192 (22 412 601 to 48 779 175) | 40 418 484 (27 096 405 to 59 175 351) | 46 666 848 (31 085 941 to 68 471 907) | 52 191 388 (34 420 729 to 76 775 591) | 56 653 481 (36 886 513 to 83 581 733) | 58 893 762 (37 941 189 to 87 078 798) | 60 539 909 (38 716 224 to 89 648 456) | 61 333 544 (39 089 885 to 90 887 354) |
| Cost of eye protection (high estimate) | 3 799 586 (2 692 439 to 5 248 636) | 10 613 370 (7 239 367 to 15 274 257) | 17 699 994 (11 968 349 to 25 701 296) | 24 994 112 (16 829 551 to 36 443 294) | 32 259 786 (21 665 514 to 47 153 203) | 39 071 202 (26 193 190 to 57 202 840) | 45 111 287 (30 049 743 to 66 189 511) | 50 451 676 (33 273 372 to 74 216 406) | 54 765 032 (35 656 963 to 80 795 676) | 56 930 636 (36 676 483 to 84 176 173) | 58 521 912 (37 425 684 to 86 660 175) | 59 289 092 (37 786 889 to 87 857 776) |
| Excess cost (high estimate) | 37 980 082 (26 025 847 to 52 950 638) | 106 836 811 (69 884 558 to 155 120 983) | 178 450 717 (115 499 331 to 261 382 165) | 252 066 043 (162 295 806 to 370 737 845) | 325 297 967 (208 754 801 to 479 650 714) | 393 859 952 (252 162 301 to 581 738 944) | 456 171 658 (289 990 315 to 675 302 953) | 513 206 941 (322 789 665 to 761 740 798) | 561 568 897 (348 516 755 to 835 943 901) | 587 697 548 (360 817 557 to 876 730 589) | 606 896 760 (369 856 894 to 906 700 804) | 616 153 013 (374 214 946 to 921 150 217) |
| Cost of surgical procedure | 313 652 684 (207 283 723 to 441 845 550) | 891 421 270 (558 100 521 to 1 306 717 159) | 1 492 324 940 (922 963 632 to 2 206 217 802) | 2 110 140 475 (1 297 198 557 to 3 132 113 857) | 2 724 857 914 (1 668 653 766 to 4 054 463 327) | 3 300 488 852 (2 015 634 108 to 4 919 208 952) | 3 836 515 282 (2 325 783 809 to 5 730 737 258) | 4 342 289 487 (2 604 374 326 to 6 502 597 438) | 4 788 434 458 (2 834 551 401 to 7 190 253 056) | 5 042 726 142 (2 954 266 897 to 7 587 201 060) | 5 229 578 499 (3 042 239 888 to 7 878 879 961) | 5 319 663 063 (3 084 653 732 to 8 019 505 894) |
| Grand total (low estimate) | 338 563 656 (224 366 633 to 476 568 706) | 961 484 406 (603 972 898 to 1 408 425 482) | 1 609 348 236 (998 778 175 to 2 377 593 336) | 2 275 437 691 (1 403 732 340 to 3 375 187 039) | 2 938 178 990 (1 805 686 654 to 4 368 945 461) | 3 558 772 643 (2 181 164 207 to 5 300 626 994) | 4 135 641 415 (2 516 135 442 to 6 173 470 211) | 4 678 772 652 (2 816 230 681 to 7 001 936 053) | 5 156 563 120 (3 063 254 901 to 7 738 140 700) | 5 427 928 489 (3 191 008 318 to 8 161 740 638) | 5 627 326 510 (3 284 888 036 to 8 473 003 481) | 5 723 459 544 (3 330 149 635 to 8 623 071 337) |
| Grand total (high estimate) | 351 632 766 (233 309 570 to 494 796 188) | 998 258 081 (627 985 079 to 1 461 838 142) | 1 670 775 657 (1 038 462 963 to 2 467 599 967) | 2 362 206 518 (1 459 494 363 to 3 502 851 702) | 3 050 155 881 (1 877 408 567 to 4 534 114 041) | 3 694 348 804 (2 267 796 409 to 5 500 947 896) | 4 292 686 940 (2 615 774 124 to 6 406 040 211) | 4 855 496 428 (2 927 163 991 to 7 264 338 236) | 5 350 003 355 (3 183 068 156 to 8 026 196 957) | 5 630 423 690 (3 315 084 454 to 8 463 931 649) | 5 836 475 259 (3 412 096 782 to 8 785 580 765) | 5 935 816 076 (3 458 868 678 to 8 940 656 111) |

**Supplementary table 10.** Cost of resources for cumulative deficit of surgical activity between 1^st^ March 2020 and 28^th^ February 2021, reported on the last day of each month. Total number of admissions is presented as a time-weighted average with 95% confidence intervals derived from a linear growth model at procedure group level. Costs are presented in pounds sterling and include only excess costs associated with specific measures required for surgical time. Low estimate: four members of staff in personal protective equipment, bed day cost of £222 (€243.68). High estimate: eight members of staff in personal protective equipment, bed day cost of £346 (€379.79). Costs are provided in Euros (€).

| **Class** | **Mar** | **April** | **May** | **Jun.** | **Jul.** | **Aug.** | **Sep.** | **Oct.** | **Nov.** | **Dec.** | **Jan.** | **Feb.** |
| --- | --- | --- | --- | --- | --- | --- | --- | --- | --- | --- | --- | --- |
| **Class 1** | 0 (0 to 0) | 0 (0 to 0) | 0 (0 to 0) | 0 (0 to 0) | 0 (0 to 0) | 0 (0 to 0) | 0 (0 to 0) | 0 (0 to 0) | 0 (0 to 0) | 0 (0 to 0) | 0 (0 to 0) | 0 (0 to 0) |
| **Class 2** | 7 058 (5 840 to 8 555) | 13 699 (11 181 to 16 912) | 20 606 (16 736 to 25 604) | 26 738 (21 667 to 33 320) | 31 860 (25 786 to 39 765) | 35 726 (28 896 to 44 630) | 38 723 (31 306 to 48 401) | 40 822 (32 994 to 51 042) | 41 866 (33 834 to 52 356) | 41 866 (33 833 to 52 356) | 41 866 (33 834 to 52 356) | 41 866 (33 834 to 52 356) |
| **Class 3** | 84 127 (69 109 to 110 982) | 226 800 (186 645 to 311 721) | 375 186 (308 887 to 520 497) | 506 915 (417 407 to 705 837) | 616 949 (508 054 to 860 652) | 699 994 (576 468 to 977 495) | 764 377 (629 508 to 1 068 081) | 809 467 (666 653 to 1 131 521) | 831 896 (685 130 to 1 163 078) | 831 896 (685 130 to 1 163 078) | 831 896 (685 130 to 1 163 078) | 831 896 (685 130 to 1 163 078) |
| **Class 4** | 58 019 (30 779 to 86 569) | 176 272 (86 453 to 271 164) | 299 260 (144 356 to 463 150) | 408 442 (195 759 to 633 584) | 499 642 (238 696 to 775 948) | 568 473 (271 102 to 883 394) | 621 836 (296 226 to 966 695) | 659 208 (313 821 to 1 025 033) | 677 798 (322 573 to 1 054 052) | 677 798 (322 573 to 1 054 052) | 677 798 (322 573 to 1 054 052) | 677 798 (322 573 to 1 054 052) |
| **Total** | 149 204 (105 728 to 206 106) | 416 771 (284 279 to 599 797) | 695 052 (469 979 to 1 009 251) | 942 095 (634 833 to 1 372 741) | 1 148 451 (772 536 to 1 676 365) | 1 304 193 (876 466 to 1 905 519) | 1 424 936 (957 040 to 2 083 177) | 1 509 497 (1 013 468 to 2 207 596) | 1 551 560 (1 041 537 to 2 269 486) | 1 551 560 (1 041 536 to 2 269 486) | 1 551 560 (1 041 537 to 2 269 486) | 1 551 560 (1 041 537 to 2 269 486) |

**Supplementary Table 11.** Cumulative deficit of surgical activity on the last day of each month from a post-hoc sensitivity analysis model for reintroduction of surgical activity between 1^st^ March 2020 and 28^th^ February 2021. Assumes: continued class 1 (emergency) activity at pre-pandemic levels; continued class 2 (urgent) activity at 80% of pre-pandemic levels and increasing on 1^st^ June to reach pre-pandemic levels by 30^th^ November; reintroduction of class 3 and 4 activity on 1^st^ June and reaching pre-pandemic levels by 30^th^ November. Numbers are presented as predicted time-weighted average with 95% confidence intervals.
